# Genetic analysis of blood molecular phenotypes reveals regulatory networks affecting complex traits: a DIRECT study

**DOI:** 10.1101/2021.03.26.21254347

**Authors:** Ana Viñuela, Andrew A. Brown, Juan Fernandez, Mun-Gwan Hong, Caroline A. Brorsson, Robert W. Koivula, David Davtian, Théo Dupuis, Ian M. Forgie, Jonathan Adam, Kristine H. Allin, Robert Caiazzo, Henna Cederberg, Federico De Masi, Petra J.M. Elders, Giuseppe N. Giordano, Mark Haid, Torben Hansen, Tue Hansen, Andrew T. Hattersley, Alison J. Heggie, Cédric Howald, Angus G. Jones, Tarja Kokkola, Markku Laakso, Anubha Mahajan, Andrea Mari, Timothy J. McDonald, Donna McEvoy, Miranda Mourby, Petra Musholt, Birgitte Nilsson, François Pattou, Deborah Penet, Violeta Raverdy, Martin Ridderstrale, Luciana Romano, Femke Rutters, Sapna Sharma, Harriet Teare, Leen M T’Hart, Kostas Tsirigos, Jagadish Vangipurapu, Henrik Vestergaard, Søren Brunak, Paul W. Franks, Gary Frost, Harald Grallert, Bernd Jablonka, Mark I. McCarthy, Imre Pavo, Oluf Pedersen, Hartmut Ruetten, Mark Walker, the DIRECT consortium, Jerzy Adamski, Jochen M. Schwenk, Ewan R. Pearson, Emmanouil T. Dermitzakis

**Author notes:** Corresponding authors: Ana Viñuela and Emmanouil T. Dermitzakis.

## Abstract

Genetic variants identified by genome-wide association studies can contribute to disease risk by altering the production and abundance of mRNA, proteins and other molecules. However, the interplay between molecular intermediaries that define the pathway from genetic variation to disease is not well understood. Here, we evaluated the shared genetic regulation of mRNA molecules, proteins and metabolites derived from whole blood from 3,029 human donors. We find abundant allelic heterogeneity, where multiple variants regulate a particular molecular phenotype, and pleiotropy, where a single variant was associated with multiple molecular phenotypes over multiple genomic regions. We find varying proportions of shared genetic regulation across phenotypes, highest between expression and proteins (66.6%). We were able to recapitulate a substantial proportion of gene expression genetic regulation in a diverse set of 44 tissues, with a median of 88% shared associations for blood expression and 22.3% for plasma proteins. Finally, the genetic and molecular associations were represented in networks including 2,828 known GWAS variants. One sub-network shows the trans relationship between rs149007767 and *RTEN*, and identifies *GRB10* and *IKZF1* as candidates mediating genes. Our work provides a roadmap to understanding molecular networks and deriving the underlying mechanism of action of GWAS variants across different molecular phenotypes.

## Introduction

Genome-wide association studies (GWAS) can explain how genetic variation contributes to phenotypic variation by associating particular genomic regions to a trait of interest, often with the underlying assumption that one or multiple genes mediate this association. However, identifying the mediating molecules is still a challenge, as a large proportion of GWAS variants are located in non-coding regions with no obvious gene target^1^. Molecular studies at the population level can be used to identify these mediating molecules, by testing whether the genetic variant associated with a complex trait is also associated with gene expression levels. These expression quantitative trait loci (eQTLs) studies have been very successful in identifying molecular regulatory regions and candidate genes^2,3^. However, to understand the full causal relationship between genetic variants and complex traits, and the various stages appropriate for clinical intervention, we require studies with deep molecular phenotyping.

Recent studies have focused on molecular regulatory processes associated to GWAS studies affecting the abundance of phenotypes other than mRNA expression, such as circulating metabolites ^4^, plasma proteins^5–7^ or other molecular phenotypes^8,9^. However, these studies often focus on one additional type of molecular phenotype, revealing only a few elements of the complete molecular path between a GWAS variant and disease. The few studies that have explored the relationships between multiple phenotypes derived from samples from the same individuals are often limited in sample size for genetics analyses ^10^. An additional challenge for these types of study is to integrate genetic associations across molecular phenotypes to understand the downstream consequences of genetic perturbations and their cascade effects through different layers of regulation. Integrated information from these studies will be key to understand how the relationships between multiple perturbations and phenotypes define complex trait variability^11^.

Here, we use data from the DIabetes REsearCh on patient straTification (DIRECT) consortium to investigate the genetic regulation of multiple molecular phenotypes. Using blood and plasma samples from DIRECT participants newly diagnosed with diabetes or consider pre-diabetics, we performed local (cis) and distal (trans) quantitative trait loci (QTLs) analysis in derived gene expression, protein and metabolites phenotypes. A deep analysis of these QTLs investigated allelic heterogeneity and pleotropic effects shared across molecular phenotypes with local and distal effects. Visualization and characterization of regulatory networks from shared QTLs across phenotypes was the used to connect clusters of genetic regulation across phenotype to GWAS signals. Our work has implications for the mechanistic understanding of the activity of GWAS variants, the identification of therapeutic targets for complex genetic diseases treatment and the general principles of genetic regulation of molecular phenotypes and complex traits.

### Identification of local genetic regulation of molecular phenotypes

The DIRECT dataset consists of 3,029 individuals of European descent including pre-diabetic participants and patients with newly diagnosed type 2 diabetes with genotype information, expression quantified by RNA sequencing (RNA-seq), targeted proteomics (multiplexed immuno-assays) and metabolites (targeted and untargeted mass spectrometry) in whole blood (Methods, Supplementary Table 1). To identify independent local (cis-) genetic effects that regulate mRNA expression (cis-eQTLs) and levels of circulating protein (cis-pQTLs), we performed a QTL analysis followed by a stepwise regression conditional analysis (Figure 1A). We identified independent significant associations for 94.4% (15,305) of genes expression and 97.3% (363) of proteins (Supplementary Table 2, Methods, Supplementary Data 1-3), finding similarities in the regulation of both expression and proteins (Supplementary Figures 1 and 2). For example, 83.8% of the genes and 86.7% of the proteins with cis-QTLs were associated with multiple SNPs, demonstrating extensive allelic heterogeneity. However, functional enrichment analysis of eSNPs relative to pSNPs, using available ChromHMM annotations from 14 blood cell lines^12^ and VEP annotations^13^, found these two classes of regulatory variants had different properties. pSNPs were enriched in 5’ UTR variants relative to eSNPs (OR=2.84, Pvalue=8.6e-16), while eSNPs were enriched in active TSS (OR E044=4.95, Pvalue=7.7e-03) (Supplementary Figure 3, Supplementary Data 4). In summary, the abundance of independent local genetic regulation identified here supports extensive allelic heterogeneity regulating molecular phenotypes^14^.

**Figure 1A.**
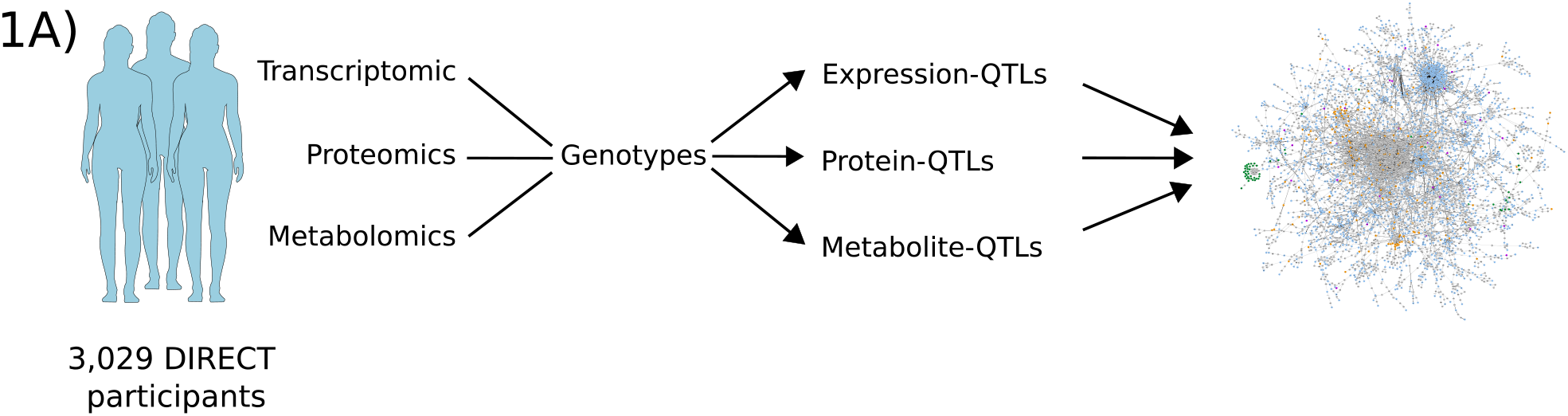
Multi-omic QTL analysis identifies extensive allelic heterogeneity and pleotropic effects. The DIRECT consortium collected blood and plasma samples from 3,029 individuals from which it derived genetic, transcriptomic, proteomic and metabolite data. The data was then used to identify genetic associations with expression (eQTLs), proteins (pQTLs) and metabolites (metabolite-QTLs). We explored the QTLs to identify shared and specific genetic regulatory effects across molecular phenotypes, and also combined associations to build a network of the genetic perturbations affecting molecular phenotypes. The network shown here corresponds to the larger cluster of connected nodes from the full network.

Pleiotropic effects, when one SNP affects multiple molecular phenotypes, were also common among cis-QTLs and identified networks of local regulatory genetic effects shared across nearby genes (Figure 1B). For cis-eQTLs, these networks included 1,924 examples of one SNPs (eSNP) associated to the expression two or more genes (Supplementary Data 5). For proteins with limited number of phenotypes (n=373), just 3 examples were identified with the SNPs associated (pSNP) to two proteins with the same direction of effect: rs2405442 regulated PILRB and PILRA, rs1130371 for CCL18 and CCL3, and rs7245416 for CD97 and EMR2 (Supplementary Figure 4). For cis-eQTLs, genes with shared eSNPs had a mean distance of 0.14 Mb, with 30.30% of the pairs showing the opposite direction of effect for the two cis-eQTLs (Supplementary Figure 5). For example, the C allele in rs907612, a variant previously associated with monocyte abundance^15^, increased expression of the lymphocyte specific protein 1 (LSP1) while decreasing the expression of IFITM10, a gene that codes for the interferon induced transmembrane protein 10 (Supplementary Figure 6). Genes with opposite effect eSNPs were more likely to be further away from each other than those with the same direction of effect (Wilcoxon test Pvalue=7.16e-15). In summary, our results support previous reports of abundant pleiotropic effects on cis-eQTLs^14^ with limited information for cis-pQTLs. Given the increased number of proteins and samples evaluated in pQTL studies and reports of overlapping genetic architecture properties with gene expression^16–18^, we expect these pleiotropic effects to be also abundant at the protein level.

**Figure 1B.**
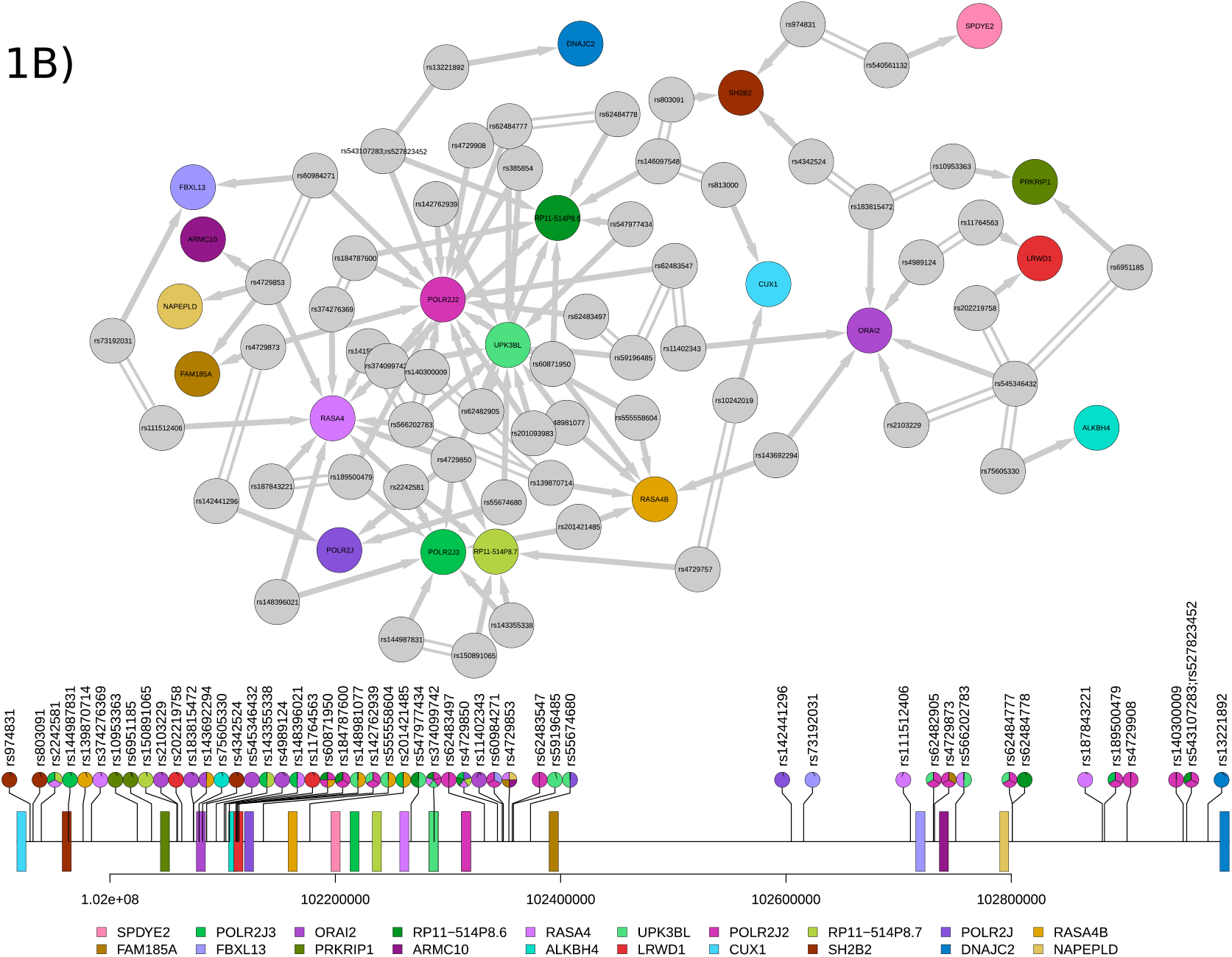
Multi-omic QTL analysis identifies extensive allelic heterogeneity and pleotropic effects. Integration of cis-eQTLs identified networks of local regulatory genetic effects. For cis-eQTLs these networks included eSNPs or pairs of eSNPs in high LD (R2>0.9) that regulated the expression of two or more genes. The upper diagram shows the network around the cis-window of POLR2J3. The lower lollipop plot shows the genomic location of the genes (boxes) and SNPs (lollipops), coloured by the associated gene.

### Distal genetic regulation shared properties with local regulation

Distal genetic regulation also exhibited allelic heterogeneity and pleotropic genetic effects for gene expression, proteins and metabolites. To identify trans-QTLs, we first performed genome-wide discovery analyses identifying 1,670 (11.04%) and 139 (37.26%) significantly associated SNPs with expression and proteins, excluding a 5Mb window around the TSS of each phenotype (Methods). Metabolites, which do not have a genomic location of reference such as a TSS, were evaluated without excluding any window, identifying 172 metabo-QTLs (49.2%). Using a conditional analysis scan we identified independently associated trans-QTLs for each of the three phenotypes. Similar to cis-QTLs, we found evidence that multiple variants were often associated with a phenotype in trans, with an average of 1.38 independent trans associations discovered for each gene, 1.18 for proteins and 1.75 for metabolites (Supplementary Table 1, Supplementary Data 5-8, Supplementary Figures 7, 8 and 9). Similarly, 20.65% of genes with at least one trans-QTL were associated with 2 or more variants, compared to 25.46% of proteins and 39.53% of metabolites. In contrast, pleiotropic regulation was less common for proteins with 8.24% of the pSNPs associated with 2 or more proteins in trans, compared to 14.57% and 17.79% of the SNPs associated with expression and metabolites in trans (Supplementary Figure 10).

Trans effects on expression have been shown to also act as cis-eQTLs^19^. For example, a local effect on the expression of a transcription factor can have downstream consequences on the expression of a distal gene. To investigate the common regulatory processes between cis and trans regulation, we looked for trans-QTLs which also acted as cis-QTLs and found significant cis-eQTL effects for 19.39% (n=262) of 1,351 trans-eSNPs (Supplementary Figure 11). For proteins, we found no trans-pSNPs that were also acting as cis-pSNPs. Given that these comparisons are limited by differences in significance thresholds and multiple testing, we estimated the proportion of significant trans-eSNPs that also affected local molecular phenotypes and applied the π1 methodology to estimate the proportion of associations from the alternative hypothesis^3,20^. We estimated that 77.34% of trans-eSNPs and 0% of trans-pSNPs had an effect on local expression and local proteins levels, respectively (Supplementary Figure 12). This estimate was higher for trans-pSNPs acting also as cis-eQTLs (91%), while there was no evidence of trans-eSNPs acting as cis-pSNPs (π1 = 0%). Next, we investigated if cis-eSNPs that also have an effect in trans were more likely to regulate a transcription factor (TF) in cis (Methods, Supplementary Data 8)^21^ and found a significant enrichment of TFs in genes for which the cis-eQTL was also associated with distal genes (OR=2.26, Pvalue=5.09e-08). In conclusion, our results support a complex interplay between local and distal regulation of the same phenotypes were trans-QTLs activity was mediated by local regulation for both expression and proteins. These distal effects were often driven by local regulation of TFs for trans-eQTLs.

### Phenotype and tissue specific genetic regulation

We next explored the degree of sharing of genetics regulation across molecular phenotypes by comparing first cis-eQTLs with cis-pQTLs. Of the 373 proteins investigated, 287 had gene expression available for the coding gene in whole blood, while the expression of 86 genes was not detected (Supplementary Data 9). For the available gene-proteins pairs, we compared the Pvalue distribution of significant cis-pQTLs with Pvalues for the same SNP-gene pair from the cis-eQTLs analysis. We observed a Pvalue enrichment of 66.58% for all pSNPs (73.87% considering only the stronger pQTLs), suggesting a large proportion of cis-eQTLs acted also as pQTLs, even when they were not individually significant. Of the significant cis-QTLs detected, 101 cases had a SNPs associated with both a protein and the expression of a gene (20.9% of the proteins). For example, rs34097845 was associated with both the expression of *MPO* and its protein (MPO) with a consistent direction of effect (Supplementary Figure 13). Of those, 53 SNPs were associated with a different gene than the coding gene of the protein. From the 48 SNPs associated to a protein and its coding gene, 4 had an opposite direction of effect for the cis-QTLs effect (Supplementary Figure 14, Supplementary Data 5). In conclusion, we estimated a large proportion of shared genetic regulation across gene expression and proteins. However, as reported by others before^5,16–18,22,23^, the genetic regulation of circulating proteins seems to involve additional protein specific regulatory processes that complicates the identification of genomic regions acting in both phenotypes.

Failure to identify shared cis-QTLs across molecular phenotypes may also be driven by tissue specific genetic regulation. For example, CCL16 is a cytokine which has a strong, replicated cis-pQTL in whole blood (lead-pSNP rs10445391, Pvalue = 9.57e-245). However, we did not discover a corresponding cis-eQTL for this gene, as the gene is not expressed in whole blood. GTEx v8^14^ reports the gene to be expressed mainly in the liver, with rs10445391 as a cis-eSNP with the same direction of effect that the blood cis-pQTL. This suggests that the cis-pQTL is the downstream consequence of gene expression regulation in liver (Figure 2A), and demonstrates the need to put single tissue molecular phenotypes into a whole organism context.

**Figure 2.**
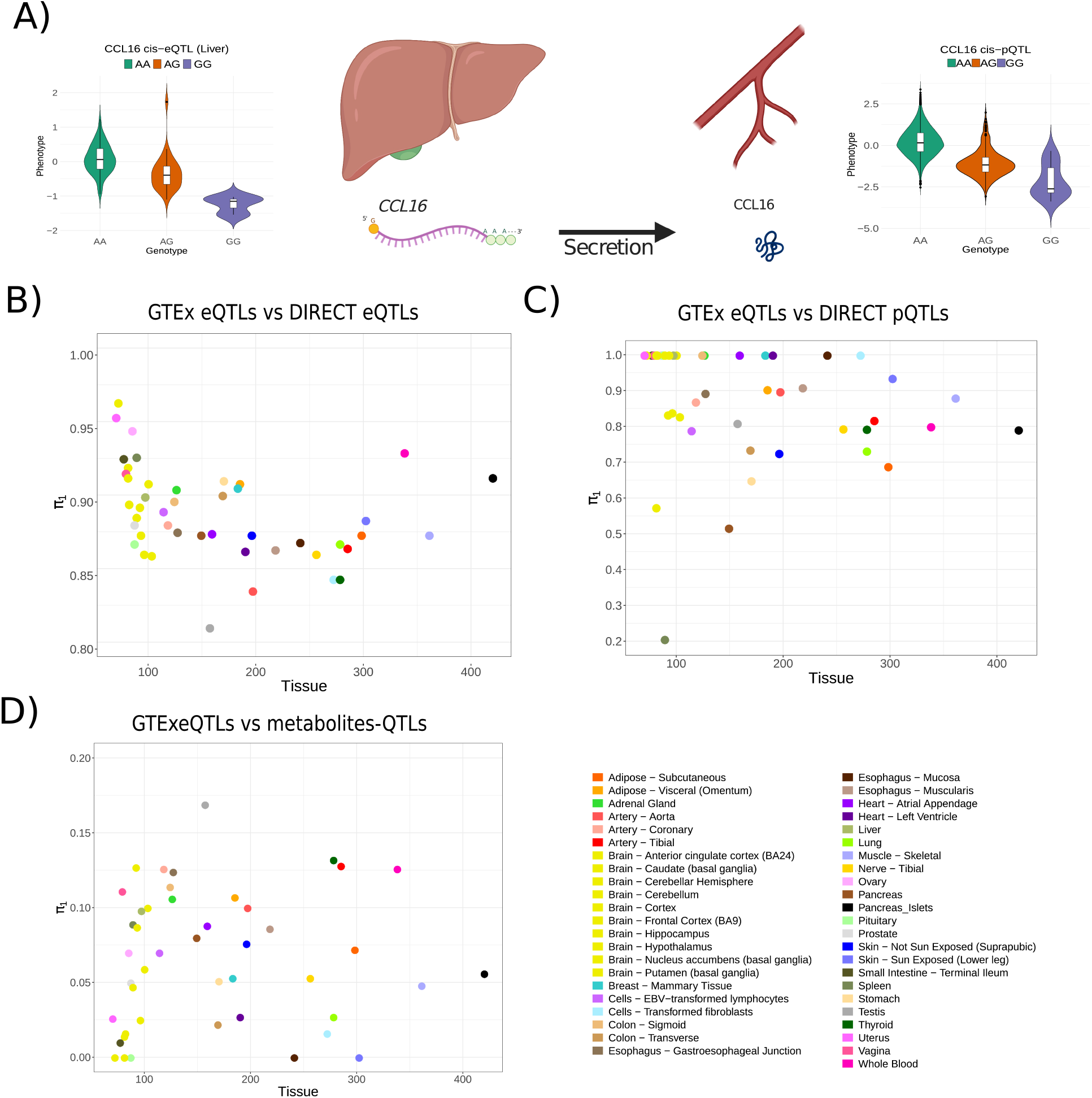
Tissue specific genetic regulation partially explains the lack of shared associations between gene expression and proteins. **A)** A cis-pQTL for CCL16 detected in whole blood is the result of genetics regulation of *CCL16* expression in liver. CCL16 is a cytokine which is mainly expressed in liver. The GTEx consortium reported cis-eQTL, with rs10445391 affecting liver expression. The protein, whose abundance is dependent on the mRNA abundance, is then released to the blood stream, where a cis-pQTL can be detected. **B)** Large transcriptional datasets in whole blood can detect the majority of genetic regulation of gene expression in other tissues. Between 81% (testis) to 96.3% (brain cortex (BA24)) of cis-eQTLs discovered by GTEx were also active in whole blood. **C)** Proteins shared a lower percentage of genetic regulatory signals with gene expression across tissues. However, 62.4% of plasma cis-pQTLs were also active whole blood cis-eQTLs. **D)** Genetic regulation of metabolites show a distinct pattern compared to gene expression and proteins. Up to 16.88% (testis) of the metabo-QTLs detected in blood were active eQTLs in other tissues, with many tissues sharing no associations with metabolites-QTLs.

To further investigate the relationship between blood and plasma molecular associations and similar processes in other tissues, we looked for evidence that cis-eQTLs detected in other tissues (43 GTEx tissues^14^ and pancreatic islets^3^ were also active in the DIRECT data from blood. Using Pvalue enrichment analysis (π1) (Methods), we compared the distribution of Pvalues for significant cis-eQTLs across different tissues in DIRECT blood eQTLs, estimating that between 81% (testis) and 96.3% (brain cortex (BA24)) of those cis-eQTLs were also active in whole blood (Figure 2B). Our results indicate that a sufficiently large sample size in blood can be informative of regulation in other tissues, although the regulation of specific genes may be missed if relevant tissues are not studied (Supplementary Figures 14-15). Next, we investigated whether these cis-eSNPs active other tissues were also regulating protein or metabolite levels from blood plasma. For cis-pQTLs, we observed π1 estimates ranging from 20.5% in spleen to 100% in multiple tissues (e.g.: liver, skeletal muscle). These estimates suggest that CCL16 is not a unique example and that genetic regulation of transcript abundance is shared with blood circulating protein levels. For metabolites with no direct match between phenotypes, we extracted all the cis-eQTLs or cis-pQTLs associated with the most significant metabo-SNPs to calculate π1 estimates. Metabo-SNPs acting also as cis-eQTLs and cis-pQTLs in DIRECT whole blood were of 33.63% and 27.03%, respectively. These estimates are lower than the cis-pQTLs detected as cis-eQTLs (66.6%) and suggest a higher degree of independent regulation across those phenotypes. Enrichment estimates for the other 44 tissues also detected a lower proportion of shared associations for metabo-QTLs than for other molecular phenotypes, with estimates ranging from 0% (brain) to 16.88% (testis) (Figure 2). Overall, our results indicate that blood gene expression and protein levels share a large degree of genetic regulation, both within and across tissues, and to a much greater degree than those shared with metabolites.

### Causal networks

Given the abundant pleiotropy observed, with single SNPs associated with multiple molecular phenotypes, we were interested in characterizing the chain of action of genetic variation on molecular phenotypes. Therefore, we tested causal networks from 65,682 trios consisting of 14,288 SNPs significantly associated with two molecular phenotypes in cis or trans. These trios are partially directed, as causation must travel outwards from the DNA (QTLs)^5,24^. We used Bayesian Networks (BN, Methods) to evaluate two main types of causative models: i) independent models, where the SNP independently regulates the two phenotypes; ii) dependent models, where the SNP regulation of one phenotype depends on the activity of the other phenotype (Figure 3A). After evaluation of the best models, we identified 23,883 trios of SNPs (or two SNPs in high LD (R2>0.9)) with evidence supporting a particular casual model (Supplementary Figures 11, 17 and 18, Supplementary Data 10). All combinations of QTLs and the causative models investigated are fully described in Supplemental Figures 19 to 22.

**Figure 3A.**
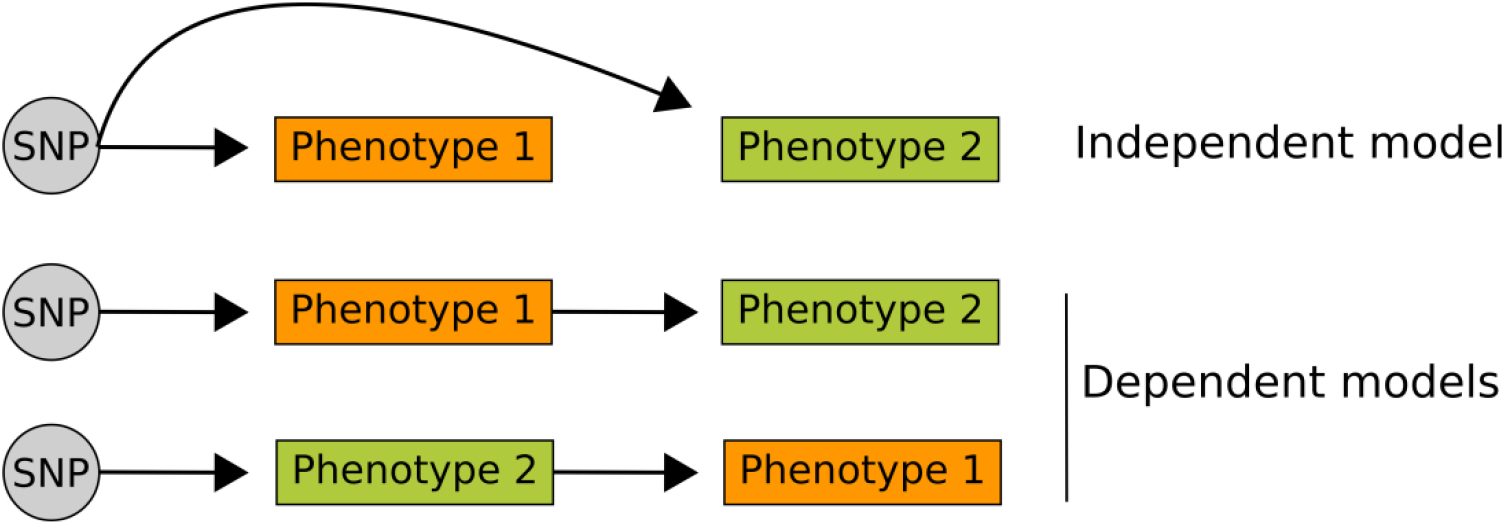
Causal inference identifies distinct patterns of casual paths in the regulation of molecular phenotypes. Two main models were tested for casual inference. The dependent model assumes the effect of a genetic variant (SNP) on phenotype 2 is mediated by phenotype 1. The independent model assumes the effect of the SNP on both SNPs is independent, and no mediation between phenotypes occurs.

**Figure 3.**
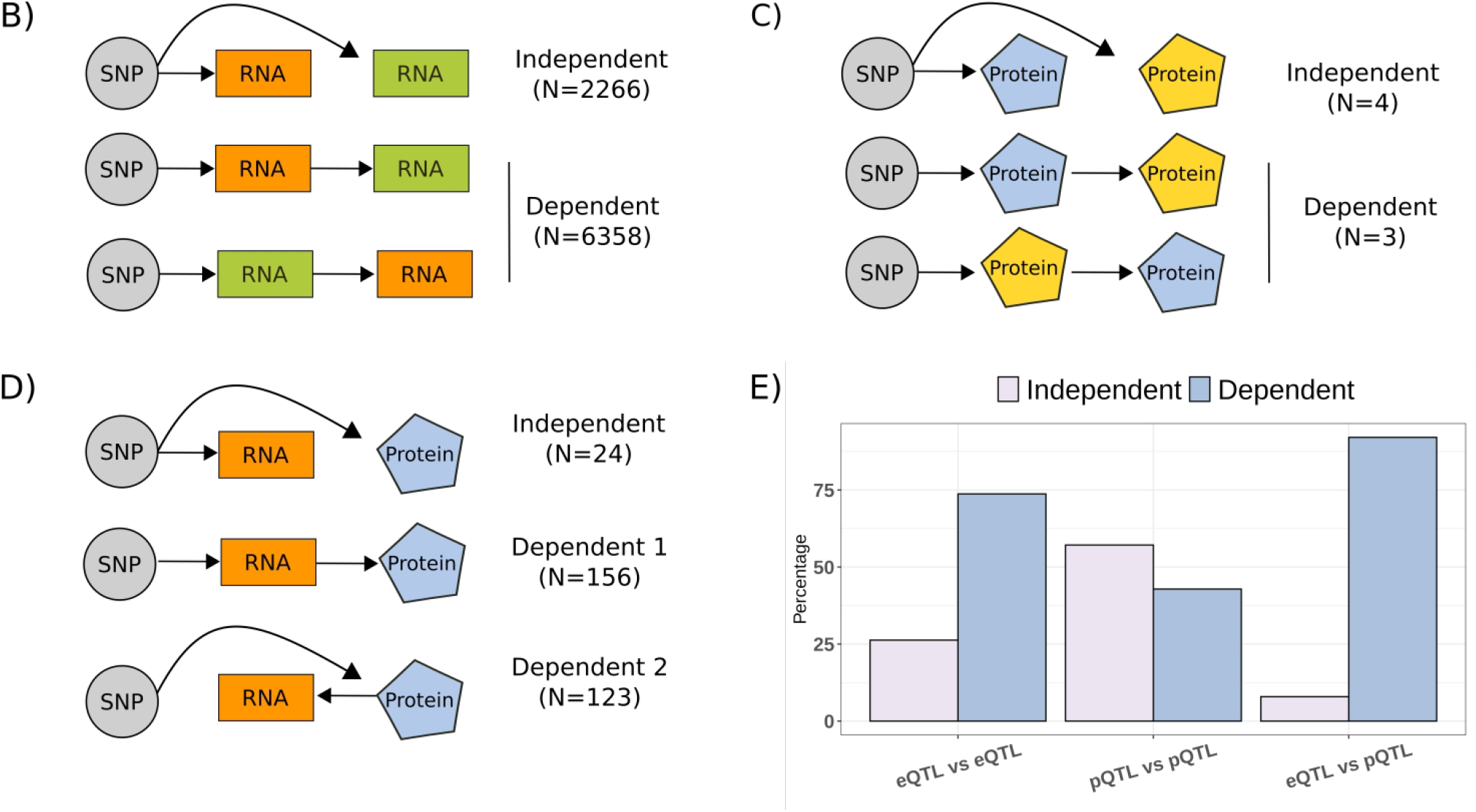
Causal inference identifies distinct patterns of casual paths in the regulation of molecular phenotypes. **B)** Casual models testing paths for SNPs acting as cis-eQTLs for two genes. We identified slightly more models supporting independent effects of the shared SNPs than dependent effects. **C)** Casual models testing paths for SNPs acting as cis-pQTLs for two proteins. Here more models supported an independent effect of the shared pSNPs. **D)** Casual models for shared QTLs between gene expression and proteins supported dependent models. When testing different molecular phenotypes, we observed a majority of cases where the evidence supported dependent models, with similar proportions where expression was the mediating factor as where the mediating factor was protein levels. **E)** The casual network analysis supports a model where the downstream consequences of genetic variation are often mediated by other molecular phenotypes.

The shared genetic regulation across different phenotypes, such as mRNA vs. proteins, had different causative relationships than the shared effects between pairs of the same molecular phenotypes, such as cis-eQTLs vs. cis-eQTLs or cis-eQTLs vs. trans-eQTLs effects. Dependent models, where the SNP’s effect on one phenotype was mediated by the expression of the other phenotype, were more often supported for trios where the SNP regulates two different molecular phenotypes (e.g.: trans-eQTL/trans-pQTL, cis-eQTL/metaboQTL) than the same molecular phenotype. For example, we found no evidence of SNPs independently affecting cis-expression and metabolites, cis-expression and trans-protein levels, or trans-expression and trans-protein levels. Only 24 out of 303 models for shared effects on cis-eQTLs and cis-pQTLs found the independent model as most likely causal (Figure 3D). An exception of this trend was observed for shared genetic regulation between two metabolites, as it was more often identified as independent (Supplementary Figure 19D). For trios involving local and distal genetics effects, we found little support for independent SNP effects. More models were supported with local associations mediating the effects on the distal phenotype when the same type of phenotype was considered (mRNA-to-mRNA), while for different phenotypes more models supported a regulation of the local phenotype mediated by the distal phenotype (Supplementary Figure 21). In conclusion, our causal network analysis supports a model where the downstream consequences of genetic variation are often mediated by other molecular phenotypes. Contra-intuitively, we also found that the path of local-distal regulation can be often mediated by distal regulation acting locally. This suggests that even well-powered studies such as this one are not yet large enough to identify the full extend of regulatory elements involved in distal regulation of molecular phenotypes.

### Integration of molecular QTLs identifies networks of GWAS variants effects

QTL studies are often used to identify candidate molecules mediating the activity of GWAS variants. However, the huge polygenicity, extensive allelic heterogeneity and pleiotropy reported for even molecular phenotypes^25^, limits our ability to identify candidate gene products, as variant’s effects can span multiple genes, assays and tissues. To simultaneously evaluate the regulatory elements of molecular phenotypes and GWAS variants, we constructed networks with nodes representing either genetic variants or molecular phenotypes and edges representing significant associations. This approach allowed us to observe allelic heterogeneity as multiple edges from genetic variants nodes pointing out to a particular molecular phenotype, and pleiotropy as multiple edges from a single genetic variant pointing out to multiple molecular phenotypes. In addition, this data representation moves beyond pairs of QTLs by adding information from GWAS studies, partially visualizing the network of GWAS variants’ effects, and their effects on mediating molecular phenotypes (Figures 1A, Supplemental Figure 23).

The complete network of all connected genetic effects on genes, proteins and metabolites included 79,733 nodes (15,254 genes, 373 proteins, 172 metabolites and 63,795 SNPs) and 80,645 edges, connected in clusters containing between 3 and 19,711 nodes. Nodes had an average of 4.31 edges connecting them to neighbouring nodes. To investigate how molecular phenotypes could have downstream consequences for the risk of disease, we extracted information from the GWAS catalogue (GWAS catalogue v1.0.2, accessed 26/10/2020^26^) and identified all SNPs that were lead GWAS variants and acted as QTLs in blood. In our network, we observed 2,828 GWAS variants (Supplementary Data 11) connected with an average to 1.9 molecular phenotypes: in total 823 genes, 58 proteins and 44 metabolites were connected to GWAS variants. First, we evaluated the level of shared signals for GWAS variants and cis-eQTLs in five studies (Methods). We found that 90.16% of blood cell counts variants and 77.04% of lipid traits variants co-localized with cis-eQTLs (COLOC probability>0.9, Methods, Supplementary Data 12). Next, using all SNPs reported as GWAS variants in the network, we found that those were connected to more molecular phenotypes than other variants in the network (Pvalue=6.7e-97, Wilcoxon test), and were more likely to be associated to proteins (OR=8.93, Pvalue=3.15e-25) or metabolites (OR=17.51, Pvalue=1e-09) than to gene expression. This enrichment could be due to GWAS variants more likely acting via processes not captured by gene expression, such as post-transcriptional modification, or it could be due to lack of statistical power. As we have more power to identify eQTLs, a higher proportion of our expression associations represent weak biological effects, with smaller downstream consequences on GWAS traits the proteins or metabolites. To evaluate the influence of statistical power for different phenotypes, we repeated the enrichment considering only the most significant expression associations, matching the number of protein associated SNPs or the number of metabolite associated SNPS (Methods). While the relative metabolite enrichment remained significant (OR=2.99, Pvalue=1.51e-3), the protein enrichment was reversed (OR=10.9, Pvalue=3.28e-81 for expression over proteins), suggesting GWAS enrichment was here not driven by post transcriptional processes. Overall, we observed that GWAS variants modulated the levels of more molecular phenotypes that non-GWAS variants associated to molecular phenotypes; in particular they were enriched in associations with metabolites and strong genetic effects on expression.

Finally, we highlight here three examples to illustrate the complexities of GWAS variant interpretation and the benefits of understanding the full regulatory context to infer their underlying mechanism of action (Figure 4). First, we evaluated the largest network cluster, with 19,711 nodes (3,362 genes, 147 proteins, 15 metabolites and 15,334 SNPs) (Figure 1A, Supplementary Figure 23). The cluster was enriched in genes and proteins involved in immune response and hematopoietic cell lineage regulation (Supplementary Data 13) and included 928 SNPs previously associated with multiple traits/diseases for blood protein levels, platelet counts, and triglycerides (Supplementary Data 13). Among the 614 trans-eSNP included in this cluster, we found a replicated trans-eQTL for rs1354034^27^; this variant was associated with the expression of 297 genes and involved in platelet function regulation^28^ (Supplemental Figure 24, Supplementary Data 5). Within this cluster, we also found the resistin gene (*RETN)*, previously associated with low-density lipoproteins (LDL) levels and cardiovascular disease^29^ (Figure 4B). *RETN* expression was associated with two trans-eSNPs: rs13284665, also associated with the expression of 16 other genes, and rs149007767, previously associated with blood cell counts^15^ and responsible for the regulation of 67 other genes. This second trans-eSNP also acted on two genes in cis, the growth factor receptor-bound protein 10 (*GRB10*) and the transcription factor IKAROS family zinc finger 1 (*IKZF1*), suggesting both would be mediating candidates genes for the trans effect on *RETN* (Supplemental Figure 25). Overall, this cluster and the observations for *RETN* highlights the level of shared genetic regulation between hematopoietic production and lipid metabolism, and demonstrates the multiple ways in which GWAS variants may modulate hundreds of molecular phenotypes.

**Figure 4A.**
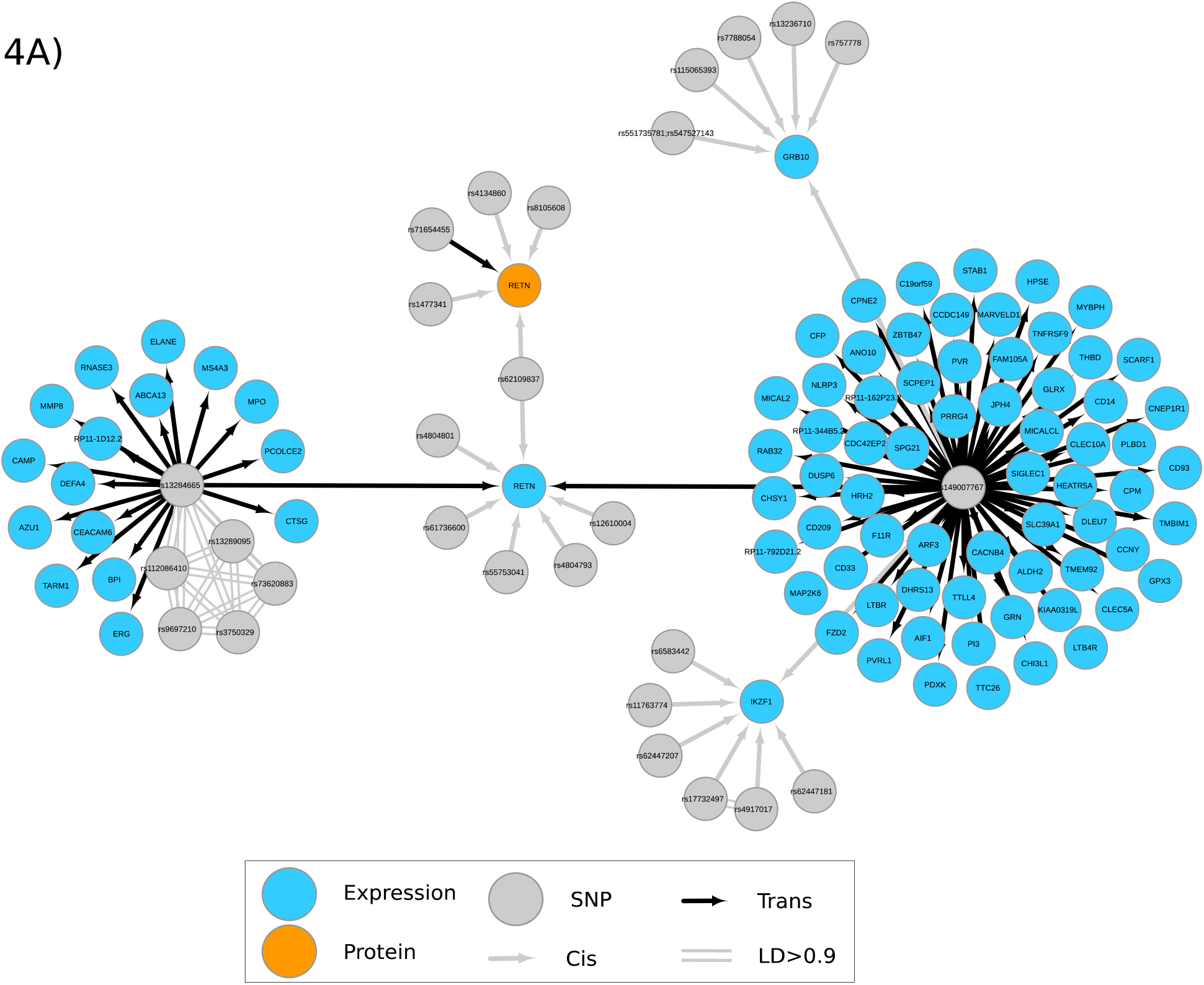
QTL integration identifies regulatory networks associated to GWAS variants. Resistin gene (*RETN)* associated network identified a genetic regulatory association between lipid metabolism and neutrophils abundance. The RETN gene and its protein (orange node) have been associated to low density lipoproteins (LDL) levels. The regulatory network associated to the gene included GWAS variants associated to cardiovascular diseases, lipid metabolism and haematopoietic processes.

**Figure 4.**
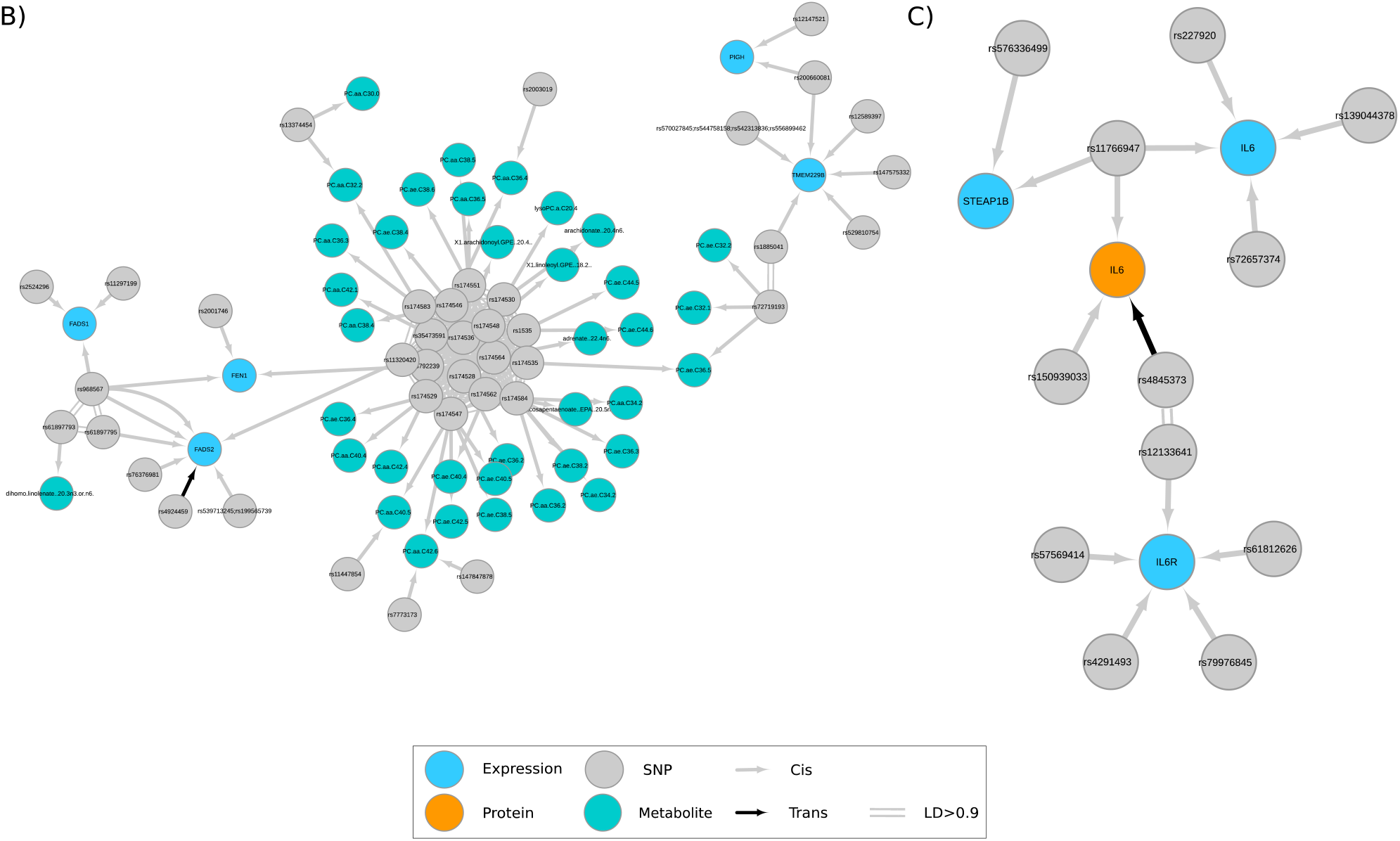
QTL integration identifies regulatory networks associated to GWAS variants. **A)** Resistin gene (*RETN)* associated network identified a genetic regulatory association between lipid metabolism and neutrophils abundance. The RETN gene and its protein (orange node) have been associated to low density lipoproteins (LDL) levels. The regulatory network associated to the gene included GWAS variants associated to cardiovascular diseases, lipid metabolism and haematopoietic processes. **B)** Network for the FADS2 gene. This network is centred in a cis-eSNP (rs968567) for FEN1, FADS1 and FADS2, which have been associated to lipid metabolism. The network shows their relationship with a cluster of genetic associations with metabolites (metabo-QTLs), many of which have been replicated by other studies. **C)** Network for the IL-6 gene. The network is centered in bot a SNP, rs11766947 associated to the expression of IL-6 and the protein levels of IL-6. We also detected a trans-pQTL associated to IL-6 in high LD with a cis-eQTL for the IL-6 receptor (IL-6R), suggesting shared regulation of both the protein and its receptor.

In a second example we focuses on the cis-window around the *FADS1* gene on chromosome 11 (Figure 4C). This region contains a cis-eSNP (rs968567) associated with *FEN1, FADS1* and *FADS2* expression previously associated with rheumatoid arthritis^30^ and fatty acid desaturase activity^31^. The region also harbours a complex locus with multiple replicated GWAS associations for metabolites^4,32^. Our results suggest multiple independent SNPs regulate fatty acid related metabolites, supporting the mediation of *FADS1*/*FADS2* in regulating plasma metabolites levels such as arachidonate (20:4n6)^33^, while also identifying other candidate effector transcripts mediating metabolites regulation such as *TMEM229B* or *FEN1*. The third example involves a shared genetic regulation of the interleukine-6 (IL-6) and its receptor (*IL-6R*) (Figure 4D). Both expression and protein levels of IL-6 were locally regulated by rs11766947 in addition to other cis-SNPs on chromosome 7. IL-6 proteins levels were also associated with a trans-pSNP on chromosome 1 in high LD (R ^2^= 0.97) with a replicated cis-eSNP (rs12133641) for the *IL-6R* gene (also known as IL-6-RA). These networks support the hypothesis that shared genetic regulation may modulate IL-6 levels, which increase after treatment with anti-IL-6 receptor antibodies for rheumatoid arthritis^17^.

Using genetic association analyses, we have evaluated the interplay between blood mRNA molecules, proteins and metabolites, as well as their genetic basis. We used genetics perturbation which allow for the orientation of effects and causal modelling to identify the regulatory principles of molecular phenotypes and potentially the genetics-to-disease paths in humans. We report large and complex cis-regulatory networks connected across different regions of the genome by trans-effects. Local allelic heterogeneity and pleotropic effects have been reported for expression and proteins^5,14,17,34–37^, but we observed here cis and trans allelic heterogeneity at larger scales than previously reported. This supports the presence of additional downstream regulation for proteins independent of gene expression regulation and provides an explanation for the reported lack of correlation between expression and protein levels^22^.

We evaluated the relevance of our findings for informing about molecular genetic regulation in other tissues. Using whole blood phenotypes, we identified a majority of the active genetics regulation of gene expression in other tissues, indicating that a sufficiently large sample size in accessible tissues can be informative of regulation in other tissues. To a lesser extent, our results show shared genetic regulation across molecular phenotypes and tissues, such as the case of secreted liver proteins (e.g.: CCL16), but also limitations in detecting tissue-specific signals, due to the lack of large studies with less-accessible tissues. Moreover, it is important to stress that true tissue specific signal and potential GWAS effector transcripts would still be missed if the causal tissue was not studied^3^.

Finally, our work provides a roadmap to understanding the underlying mechanisms of action of GWAS variants. Using networks derived from thousands of QTLs, we observed the intricate connections across molecular components regulated throughout the whole genome, rather than a single direct pathway from genotypes to phenotypes. Our results support models of genetic regulation that consider thousands of small and coordinated genetic regulatory effects across the genome to modulate complex traits^25,38–40^. Moreover, our results suggest that the regulatory process that connects genotype to phenotypes is robust, with redundancies in the form of many connections and the ability to find alternative routes in the event of a particular process being altered by disease-related variants. Therefore, as proposed by others ^41^, we have observed how a network of variants and its connected molecules as a whole is required to define a given phenotype or disease.

## Methods

### Cohort

The DIRECT (Diabetes Research on Patient Stratification) consortium includes pre-diabetic participants (target sample size 2,200–2,700) and patients with newly diagnosed type 2 diabetes (target sample size ∼1,000) with detailed metabolic phenotyping. Characteristic of the cohort as well as inclusion/exclusion criteria have been described elsewhere^42^. Fasting blood samples from venous blood were collected and DNA extractions and other biochemical analyses were carried out.

### Genotyping

Genotyping was conducted using the Illumina HumanCore array (HCE24 v1.0) and genotypes were called using Illumina’s GenCall algorithm. Samples were excluded for any of the following reasons: call rate <97%; low or excess mean heterozygosity; gender discordance; and monozygosity. Genotyping quality control was then performed to provide high-quality genotype data for downstream analyses using the following criteria: call rate <99%; deviation from Hardy-Weinberg equilibrium (exact p<0.001); variants not mapped to human genome build GRCh37; and variants with duplicate chromosome positions. To identify possible ethnical outliers in the DIRECT data, we performed a principal component analysis (PCA) using the genotype data from our studied population (3,102 samples; 547,644 markers) using the following cut-offs MAF>0.01, HWE>1e-4 and call rate>90%. A total of 3,033 samples and 517,958 markers across the two studies passed quality control procedures. Imputation to the 1000 Genomes Phase 3 CEU reference panel was performed with ShapeIt (v2.r790)^43^ and Impute2 (v2.3.2)^44^.

### RNAseq data generation

mRNA samples were processed and quality check was assessed using the TapeStation Software (A.01.04) with an RNA Screen Tape from Agilent. Quality of the libraries was evaluated using Qubit and TapeStation using DNA1000 Screen Tape. The samples were then sequenced on the Illumina HiSec2000 platform using 49 bp paired-end reads. The 49-bp sequenced paired-end reads were mapped to the GRCh37 reference genome^45^ with GEMTools 1.7.1^46^. Exon quantifications were calculated for all elements annotated on GENCODE v19^47^. Gene quantifications were calculated as FPKM values. This pipeline is fully described in Delaneau et al.^48^, as part of QTLtools. Samples with a total number of exonic reads lower than 5e+06 reads or with a proportion of exonic reads over the total number of reads lower than 20% were considered of low quality and removed. For each samples, we evaluated possible samples mix-ups^49^ using the function *match* from the suite QTLtools^50^. To confirm the correct assignment of the matched DNA/RNA samples and recovered failed genotypes during QC we re-genotyped samples from 96 individuals. Further validation compared the sex information provided by clinical reports with both genotype data and RNAseq data. The total number of samples with RNAseq-Genotypes pairing data after QC of both RNAseq and imputed genotypes was 3,029.

### Expression phenotypes

Genes and exons with more than 50% of zero reads were removed from the study. Finally, exons and genes from chromosome Y, mitochondria, and level 3 annotations, as defined by Gencode v19, were removed from further analysis. The final number of genes and exons used for analyses were 16,205 and 170,198, receptively. Splicing phenotypes were generated using LeafCutter^51^ requiring a minimum of 50 reads per cluster. Clusters with more than 50% zero reads across samples were removed. The final number of phenotypes used for further analyses were 64,546.

### Proteins data

Plasma proteins were measured using the Olink® Cardiometabolic, Cardiovascular II, Cardiovascular III, Development and Metabolism panels (Olink Proteomics AB, Uppsala, Sweden) according to the manufacturer’s instructions. The Proximity Extension Assay (PEA) technology used for the Olink protocol has been well described^52^. The obtained data was processed using Olink’s NPX manager software version 0.0.85.0. Internal and external controls were used for quality control and normalization of the data. Quality control included calculating the standard deviation for the detection control and the incubation/immuno controls and comparison of the results for the detection control and one of the incubation controls against the run median. All protein measurements were reported, but proteins with more than 50% samples bellow LOD levels were excluded, leaving a total of 373 proteins for further analysis from 3,027 individuals.

### Metabolites data

Metabolites abundance was assessed using targeted and untargeted technologies. The assays, quality control and analyses were performed separately, and results combined for discussion. Plasma targeted metabolites were evaluated for 163 metabolites using a FIA-ESI-MS/MS-based targeted metabolomics approach with the Absolute*IDQ*™ p150 kit (BIOCRATES Life Sciences AG, Innsbruck, Austria) as described in Biocrates manual AS-P150. Mass spectrometric analyses were done on an API 4000 triple quadrupole system (Sciex Deutschland GmbH, Darmstadt, Germany) equipped with a 1200 Series HPLC (Agilent Technologies Deutschland GmbH, Böblingen, Germany) and a HTC PAL auto sampler (CTC Analytics, Zwingen, Switzerland) controlled by the software Analyst 1.6.2. Data evaluation for quantification of metabolite concentrations and quality assessment was performed with the software MultiQuant 3.0.1 (Sciex) and the Met*IDQ*™ software package, which is an integral part of the Absolute*IDQ*™ Kit. Five aliquots of a pooled reference plasma were analyzed on each kit plate and used for normalization purposes and for calculation of coefficient of variance (CV) for each metabolite. Quality assessment evaluated peak shapes, retention times, compound identity, and the number of samples with zero values in the metabolites concentration, removing any individual with more than 50% of zeros. We then evaluate the CV per metabolite removing samples with CV > 0.25 relative to the reference samples. Metabolites with concentration bellow the LOD were discarded. Of the 163 metabolites, 116 passed all quality controls in 3,029 individuals.

Untargeted metabolites from human plasma samples were assessed using the Metabolon platform. Controls included a pooled matrix sample generated by taking a small volume of each experimental sample served as a technical replicate throughout the data set. Experimental samples were randomized across the platform run with QC samples spaced evenly among the injections. Metabolites concentration was assessed using Liquid Chromatography-Tandem Mass Spectrometry (LC-MS/MS). The LC-MS portion of the platform was based on a Waters ACQUITY ultra-performance liquid chromatography (UPLC) and a Thermo-Finnigan LTQ mass spectrometer operated at nominal mass resolution, which consisted of an electrospray ionization (ESI) source and linear ion-trap (LIT) mass analyzer. Raw data was extracted, peak-identified and QC processed using Metabolon’s hardware and software. For studies spanning multiple days, a data normalization step was performed to correct variation resulting from instrument inter-day tuning differences. For every metabolite we compute the coefficient of variation (CV) of measurements by run day. We then take the median CV over run days and use that as a measure of the variability of the measurement process. Metabolites with a median CV greater than 0.25 or those were the CV could not be computed for at least two run were excluded. In total we analysed 233 untargeted metabolites from 3,029 individuals.

### Cis-QTLs discovery

Local SNP-phenotypes associations were performed for gene (FPKM), exon, splice phenotypes and protein levels using linear regression in FastQTL^53^, with the seed number 1461167480. Principal component analysis (PCA) was used to control for unwanted technical variation. In addition all analyses included sex, 3 PCs derived from genotype data, and a variable identifying the cohort of origin of the samples, called “center”. Only SNPs in the near 1MB up- and down-stream of the TSS of each phenotype were considered for cis-QTLs. Raw read counts from exons and genes were used for cis-eQTLs after rank normalization, including 60 PCs for gene expression and 55 PCs for exon level. Splicing-QTLs used rank normalized phenotypes generated with LeafCutter^51^ and 20 PCs. Cis-pQTLs used rank normalize protein levels and 10PCs. Missing protein measures, no more than 8.5% for any protein, were imputed using the mean of each protein. The coding genes for the proteins was used to center cis-window for analysis. Two protein identifiers matched two genes: FUT3 and FUT5. For simplicity, these were considered independent measurements during analyses.

Multiple testing correction were performed using a beta approximation coded in FastQTL. For exon-QTLs and splicing-QTLs, we employed the grouping strategy described in Delaneau et al. (*--grp* option)^48^ to control for multiple phenotypes associated to the same TSS. This strategy computes from all phenotype-SNPs pairs in a cis-window for a given gene at once, calculating a beta distribution per gene/TSS to assess their significance. The best phenotype-SNP per gene were reported as outcome in all analyses. To control the genome-wide false discovery rate (FDR), we used the qvalue^20^ correction implemented in the software largeQvalue^54^

### Independent QTLs

Identification of secondary independent cis-QTLs was performed as described in Aguet et al^55^ and Viñuela et al.^3^using a stepwise regression procedure over all variants in the window using fastQTL, fitting all other discovered signals as covariates in addition to the other covariates and PCs. This was done only on phenotypes with an QTL discovered in the association analysis (FDR < 1%). The maximum beta-adjusted Pvalue (correcting for multiple testing across the SNPs and phenotypes) over these phenotypes was taken as the gene- or protein-level threshold. A cis-scan of the window was performed in each iteration using 1,000 permutations and correcting for all previously discovered SNPs. If this Pvalue was significant, the best association was added to the list of discovered QTLs as an independent signal and the forward step proceeded to the next iteration. The backward stage consisted of testing each forward signal separately, controlling for all other discovered signals and covariates. The exon and splicing level cis-QTL scans used the -grp function and reported only the best association in each iteration.

### Trans-QTL discovery

Trans-QTL analysis was performed between molecular phenotypes with a genomic location (RPKM expression and proteins) and all SNPs further than 5MB from the TSS of the expressed gene or coding gene for the protein. For all associations, since phenotypes PCs may capture global trans effects removing true signals, we used residuals after removing known technical covariates with a linear mixed models implemented in the lme4 package in R^56^. To control for false positives in trans-eQTLs^57^ we removed: any gene with low mappability, SNPs located in repetitive regions and with mapping issues. We then performed a genome wide scan of SNP-gene associations using QTLtools^48^ storing all Pvalues <1e-04. Pairs of SNP-genes with known cross-mapability issues in a 100kb window were then removed. Multiple testing corrections for trans-eQTLs was done using 50 permutations. The best Pvalues per phenotype were used to define a phenotype level threshold and calculate adjusted Pvalues for each gene. To adjust for multiple testing across genes, we used the qvalue package to estimate the false discovery rate. For proteins, 100 permutations per protein were used. For the identification of secondary trans-QTL, we used a stepwise regression analysis after defining non-overlapping cis-windows around 1MB up- and down-stream of any significant trans-QTLs signal. This created 2,021 genomic regions for expression and 2,589 regions for proteins to be tested for any nearby signal. Then, we run the standard conditional analysis using FastQTL to look for further signals.

### Functional annotation

Functional enrichment analysis of eSNPs relative to pSNPs was done using available ChromHMM annotations from 14 blood cell lines^12^ and VEP annotations^13^. Odds ratios for enrichment of cis-eQTLs relative to cis-pQTLs and cis-QTLs relative to trans-QTLs were computed in R.

### Transcription factor enrichment

To estimate the proportion of significant trans-SNPs which also affected local molecular phenotypes, we extracted all the cis-QTL Pvalues for SNPs with a significant trans-SNP effect and calculated π_1_ estimates using all significant associations and the qvalue package. The product of all Pvalues per SNPs we used to calculate the probability of one gene to be associated to the trans-SNP, allowing to calculate one Pvalue per phenotype using π_1_ enrichment. To investigated if cis-SNPs that also have an effect in trans were more likely to regulate a transcription factor (TF) in cis, we extracted all genes or proteins reported by Lambert et al.^21^ as transcription factors (TFs). We then used a Fisher exact test to calculate the OR of cis-eQTL affecting TFs, given the number of TFs in the whole dataset that have trans associations.

### Metabolites-QTLs

Targeted and untargeted metabolites were residualised removing technical covariates using a linear mixed model. We performed a genome-wide association for each metabolite with no exclusion of any genomic region. Multiple testing correction was performed using 100 permutations per metabolite. Secondary signals identification used stepwise regression analysis after the identification of regions around primary associations.

### Tissue and phenotype specific associations

To identify the proportion of cis-eQTLs, cis-pQTLs and metabo-QTLs active in other tissues, we extracted the same SNP-gene in GTEx v6p^55^ and InsPIRE^3^ that were significantly associated in DIRECT. To identify the proportion of eQTLs that were identified in DIRECT as QTLs of any type, we extracted the SNP-gene pairs from the significant GTEx and Inspire significant associations. Pvalues were then evaluated for enrichment using qvalue. For metabolites-QTLs with no TSS of reference, we used all cis-eQTLs in 45 studies and cis-pQTLs in DIRECT associated to the lead metabo-SNPs to calculate pi1 estimates.

### Causal inference

To identify pairs of QTLs, we selected all SNPs, or pairs of SNPs in high linkage disequilibrium (R^2^>0.9), which were significantly associated with two phenotypes in cis or trans. We identified 23,539 cases with one genetic variant and two molecular phenotypes (simple trios), independently of the genomic location of the two phenotypes. To avoid bias due to quantification correlations, we removed all pairs involving expression phenotypes where at least one exon overlaps both genes. We then used Bayesian Networks (BNs) to learn the causal relationship between pairs of QTLs. We only considered network topologies that assume a causal effect from genetic variants towards molecular phenotypes, as the opposite effect does not have biological meaning. The models evaluated were three (Figure 3): 1) Direction 1 model ([1] and [2]): The genetic variant affects first phenotype 1, then phenotype 2; 2) Direction 2 model ([3] and [4]): The genetic variant affects first phenotype 2, then phenotype 1; 3) Independent model ([5] and [6]): The genetic variant affects both phenotypes 1 and 2 independently. The probabilities can be described in the following formulas, for one or two SNPs:

1. P(Phenotype_2_|Phenotype_1_) P(Phenotype_1_|SNP) P(Phenotype_2_) P(SNP)
2. P(Phenotype_1_|Phenotype_2_) P(Phenotype_2_|SNP_2_) P(SNP_2_) P(SNP_1_)
3. P(Phenotype_1_ |Phenotype_2_) P(Phenotype_2_|SNP) P(Phenotype_1_) P(SNP)
4. Direction 2: P(Phenotype_2_|Phenotype_1_) P(Phenotype_1_|SNP_1_) P(Phenotype_2_) P(SNP_2_)
5. P(Phenotype_1_ |SNP) P(Phenotype_2_|SNP) P(SNP)
6. Independent: P(Phenotype_1_|SNP_1_) P(Phenotype_2_|SNP_2_) P(SNP_1_) P(SNP_2_)

We used normalized quantifications after removing any technical covariates. In addition, we removed from the phenotypes the effects of other QTLs by including as covariates, creating two phenotypes per trio. Computation to generate these pseudo-phenotypes were done using the –single-signal option from CaVEMaN^58^. The phenotypes were used to compute the likelihood of the 3 possible BN topologies using the R/bnlearn package^59^. The best fit was evaluated using BIC score, with a trio considered for further evaluation if one of the three models had BIC score >10 compared to each of the other two models.

### GWAS variants identification and colocalization

To identify genetics variants with known GWAS associations we used the GWAS catalogue v1.0.2, accessed 26/10/2020^26^. All SNPs with a QTL effect that were also lead GWAS variants by the catalogue were reported and used in further analysis. To provide a measure of the co-localization signals, we calculate the probability that a GWAS hit shares the same causal variant as a cis-eQTL we performed bayesian colocalisation analyses as implemented by COLOC ^60^ and in a subset of GWAS studies. We used GWAS summary statistics from two blood related traits: platelet count and lymphocyte count^15^; and three lipid related traits: LDL(Klimentidis et al. 2020), and HDL cholesterol and triglycerides^62^. SNPs were filtered to keep variants with Pvalues < 0.1 including all the variants in a 20 kb window around the lead cis-eQTL variant. The minor allele frequencies used for the analysis were those from the GWAS summary statistics. As cis-eQTL variants selected for theses analyses were previously found to be associated with the traits studied, we reported probability that both GWAS and cis-eQTLs were shared as P(H4’) = P(H4) / (P(H3) + P(H4)). Being H3 the probability of both traits to have different causal variants and H4 the probability of both traits sharing the same causal variant. For GWAS studies reporting different sample sizes per significant variant due to population differences, we calculated propabilities using the median number of samples across studies. To identify enrichment of GWAS associations among QTLs, we calculated and evaluated odd ratios using a fisher test. Since the GWAS catalog includes results from pQTLs and metabolite-QTLs studies, we removed any GWAS-SNP from blood proteins levels, metabolites levels and related phenotypes. The complete list of variants, SNPs and studies considered is included in Supplementary Data 14. To evaluate if the enrichments were due to statistical power, we evaluated the odd ratios using two sets of randomly selected SNPs from the most significant associations eSNPs matching the number of pSNP (N = 2027) and metabolite-SNPs (n=236) being evaluated.

### Networks construction

Networks were visualised using Cytoscape v3.7.2^63^. From all the QTLs results, a table was created including phenotype (target note), SNPs (source node) and type of association (interaction). The STRING enrichment function was used to evaluate the biological enrichment of some networks.

### Figures

Data and results figures were generated using R or Cytoscape^63^ v3.7.2 for the networks. Figure 2A was created with BioRender.com. Figure 1B was created using a network from Cytoscape and a lolliplot figure generated using the lolliplot() function from the trackViewer package^64^. Figures were combined in panels using Inkscape v1.0.1. Tables and objects to load networks into Cytoscape are included in the Zenodo submission (DOI: 10.5281/zenodo.4475681).

## Data Availability

Due to the type of consent provided by study participants and the ethical approvals for this study, individual-level clinical and omics data from IMI-DIRECT cohorts cannot be transferred from the centralized IMI-DIRECT repository. Requests for access to IMI-DIRECT data, including data presented here, can be made to. Requestors will be provided with information and assistance on how data can be accessed via the DIRECT Computerome secure analysis platform following submission of appropriate documentation. The IMI-DIRECT data access policy is available from www.direct-diabetes.org.
Complete summary statistics including cis and trans genetic associations are freely available in the
following DOI: 10.5281/zenodo.4475681

## Availability of data and materials

Due to the type of consent provided by study participants and the ethical approvals for this study, individual-level clinical and omics data from IMI-DIRECT cohorts cannot be transferred from the centralized IMI-DIRECT repository. Requests for access to IMI-DIRECT data, including data presented here, can be made to. Requestors will be provided with information and assistance on how data can be accessed via the DIRECT Computerome secure analysis platform following submission of appropriate documentation. The IMI-DIRECT data access policy is available from www.direct-diabetes.org.

Complete summary statistics including cis and trans genetic associations are freely available in the following DOI: 10.5281/zenodo.4475681

## Ethics statements

Approval for the study protocol was obtained from each of the regional research ethics review boards separately (Lund, Sweden: 20130312105459927, Copenhagen, Denmark: H-1-2012-166 and H-1-2012-100, Amsterdam, Netherlands: NL40099.029.12, Newcastle, Dundee and Exeter, UK: 12/NE/0132), and all participants provided written informed consent at enrolment. The research conformed to the ethical principles for medical research involving human participants outlined in the Declaration of Helsinki.

## Acknowledgments

The work leading to this publication has received support from the Innovative Medicines Initiative Joint Undertaking under grant agreement n°115317 (DIRECT, http://www.direct-diabetes.org/), resources of which are composed of financial contribution from the European Union’s Seventh Framework Programme (FP7/2007-2013) and EFPIA companies’ in kind contribution.

## Authors contribution

Conceptualization: AV, AAB, and ETD. Methodology: AV, AAB. Software: AV, AAB. Validation: AV. Formal analysis: AV, DD, TD. Investigation: AV, RC, MH, CH, AGJ, TK, Anu.M, And.M, TJM, FP, DP, VR, LR, FR, PWF, GF, MW, JA, JMS. Resources: KHL, RC, HC, PJME, MH, To.H, Tu.H, ATH, AJH, ML, And.M, TJM, DM. MM, PM, FP, VR, MR, FR, HT, LT, JV, HV, OP. Data curation: AV, JF, MGH, CAB, RWK, RA, FD, MH, CH, Anu.M, And.M, TJM, BN, FP, VR, FR, SS, KT. Writing-original draft: AV, AAB. Writing-review & editing: AV, AAB, GN, FR, JMS, ERP, ETD. Visualization: AV, TD, DD. Supervision: BJ, ER, ETD. Project administration: AV, JF, CAB, RWK, IMF, GN, HG, BJ, ERP. Funding acquisition: SB, PWF, GF, HG, BJ, MIM, IP, HR, MW, JA, JMS, ERP, ETD.

## List of Supplementary Documents

1. Main Figures and legends are included also in a separate document.
2. Supplementary Datasets:
  1. List of 59,972 independent cis-eQTLs.
  2. List of 1,592 independent cis-pQTLs.
  3. List of 3,978 independent splicing-QTLs.
  4. Functional enrichment results.
  5. List of all SNPs associated with two phenotypes and/or in cis and trans (n=69,532 trios).
  6. List of 2,320 independent trans-eQTLs.
  7. List of 533 independent trans-pQTLs.
  8. List of 301 independent metabolites-QTLs.
  9. List of pQTLs and the matching eQTLs.
  10. Causal model that passed evaluation criteria.
  11. List of 3,652 QTLs with a GWAS SNPs.
  12. Results from the co-localization analysis analysis.
  13. STRING enrichment analysis output from the big cluster network from Cytoscape.
  14. List of excluded GWAS for enrichment analysis
  15. DIRECT consortium full list
3. Supplementary Information document:
  ‐ Supplementary tables.
  ‐ Supplementary Figures.

## References

1. Mahajan, A. et al. Fine-mapping type 2 diabetes loci to single-variant resolution using high-density imputation and islet-specific epigenome maps. Nat Genet 50, 1505–1513 (2018).

2. Gamazon, E. R. et al. Using an atlas of gene regulation across 44 human tissues to inform complex disease-and trait-associated variation. Nature Genetics 50, 956–967 (2018).

3. Viñuela, A. et al. Genetic variant effects on gene expression in human pancreatic islets and their implications for T2D. Nature Communications 11, 4912 (2020).

4. Shin, S.-Y. et al. An atlas of genetic influences on human blood metabolites. Nature Genetics 46, 543– 550 (2014).

5. Chick, J. M. et al. Defining the consequences of genetic variation on a proteome-wide scale. Nature 534, 500–505 (2016).

6. Yao, C. et al. Genome-wide mapping of plasma protein QTLs identifies putatively causal genes and pathways for cardiovascular disease. Nat Commun 9, 3268 (2018).

7. Folkersen, L. et al. Mapping of 79 loci for 83 plasma protein biomarkers in cardiovascular disease. PLOS Genetics 13, e1006706 (2017).

8. Gutierrez-Arcelus, M. et al. Passive and active DNA methylation and the interplay with genetic variation in gene regulation. eLife 2, e00523 (2013).

9. Grundberg, E. et al. Mapping cis-and trans-regulatory effects across multiple tissues in twins. Nat Genet 44, 1084–1089 (2012).

10. Zhou, W. et al. Longitudinal multi-omics of host–microbe dynamics in prediabetes. Nature 569, 663– 671 (2019).

11. Ye, Y., Zhang, Z., Liu, Y., Diao, L. & Han, L. A Multi-Omics Perspective of Quantitative Trait Loci in Precision Medicine. Trends in Genetics 36, 318–336 (2020).

12. Ernst, J. & Kellis, M. ChromHMM: automating chromatin-state discovery and characterization. Nat Methods 9, 215–216 (2012).

13. McLaren, W. et al. The Ensembl Variant Effect Predictor. Genome Biology 17, 122 (2016).

14. Aguet, F. & et al. The GTEx Consortium atlas of genetic regulatory effects across human tissues. Science 369, 1318–1330 (2020).

15. Astle, W. J. et al. The Allelic Landscape of Human Blood Cell Trait Variation and Links to Common Complex Disease. Cell 167, 1415-1429.e19 (2016).

16. Sun, B. B. et al. Genomic atlas of the human plasma proteome. Nature 558, 73–79 (2018).

17. Folkersen, L. et al. Genomic and drug target evaluation of 90 cardiovascular proteins in 30,931 individuals. Nature Metabolism (2020) doi:10.1038/s42255-020-00287-2.

18. Zhang, J. et al. Large Bi-Ethnic Study of Plasma Proteome Leads to Comprehensive Mapping of cis-pQTL and Models for Proteome-wide Association Studies. bioRxiv 2021.03.15.435533 (2021) doi:10.1101/2021.03.15.435533.

19. Bryois, J. et al. Cis and Trans Effects of Human Genomic Variants on Gene Expression. PLOS Genetics 10, e1004461 (2014).

20. Storey, J. D. A direct approach to false discovery rates. Journal of the Royal Statistical Society: Series B (Statistical Methodology) 64, 479–498 (2002).

21. Lambert, S. A. et al. The Human Transcription Factors. Cell 172, 650–665 (2018).

22. Robins, C. et al. Genetic control of the human brain proteome. http://biorxiv.org/lookup/doi/10.1101/816652 (2019) doi:10.1101/816652.

23. Mirauta, B. A. et al. Population-scale proteome variation in human induced pluripotent stem cells. eLife 9, e57390 (2020).

24. Schadt, E. E. et al. An integrative genomics approach to infer causal associations between gene expression and disease. Nat Genet 37, 710–717 (2005).

25. Sinnott-Armstrong, N., Naqvi, S., Rivas, M. & Pritchard, J. K. GWAS of three molecular traits highlights core genes and pathways alongside a highly polygenic background. eLife 10, e58615 (2021).

26. Buniello, A. et al. The NHGRI-EBI GWAS Catalog of published genome-wide association studies, targeted arrays and summary statistics 2019. Nucleic Acids Res 47, D1005–D1012 (2019).

27. Võsa, U. et al. Unraveling the polygenic architecture of complex traits using blood eQTL metaanalysis. http://biorxiv.org/lookup/doi/10.1101/447367 (2018) doi:10.1101/447367.

28. Nath, A. P. et al. An interaction map of circulating metabolites, immune gene networks, and their genetic regulation. Genome Biology 18, 146 (2017).

29. Ding, Q., White, S. P., Ling, C. & Zhou, W. Resistin and Cardiovascular Disease. Trends in Cardiovascular Medicine 21, 20–27 (2011).

30. Okada, Y. et al. Genetics of rheumatoid arthritis contributes to biology and drug discovery. Nature 506, 376–381 (2014).

31. Marklund, M. et al. Genome-Wide Association Studies of Estimated Fatty Acid Desaturase Activity in Serum and Adipose Tissue in Elderly Individuals: Associations with Insulin Sensitivity. Nutrients 10, 1791 (2018).

32. Draisma, H. H. M. et al. Genome-wide association study identifies novel genetic variants contributing to variation in blood metabolite levels. Nat Commun 6, 7208 (2015).

33. Ndungu, A., Payne, A., Torres, J. M., van de Bunt, M. & McCarthy, M. I. A Multi-tissue Transcriptome Analysis of Human Metabolites Guides Interpretability of Associations Based on Multi-SNP Models for Gene Expression. Am J Hum Genet 106, 188–201 (2020).

34. Liu, Y. et al. Quantitative variability of 342 plasma proteins in a human twin population. Molecular Systems Biology 11, 786 (2015).

35. Sun, W. et al. Common Genetic Polymorphisms Influence Blood Biomarker Measurements in COPD. PLoS Genet 12, e1006011 (2016).

36. Gallois, A. et al. A comprehensive study of metabolite genetics reveals strong pleiotropy and heterogeneity across time and context. Nat Commun 10, 1–13 (2019).

37. Wang, Y. et al. Comprehensive Cis-Regulation Analysis of Genetic Variants in Human Lymphoblastoid Cell Lines. Front. Genet. 10, (2019).

38. Boyle, E. A., Li, Y. I. & Pritchard, J. K. An Expanded View of Complex Traits: From Polygenic to Omnigenic. Cell 169, 1177–1186 (2017).

39. Chakravarti, A. & Turner, T. N. Revealing rate-limiting steps in complex disease biology: The crucial importance of studying rare, extreme-phenotype families. BioEssays 38, 578–586 (2016).

40. Civelek, M. & Lusis, A. J. Systems genetics approaches to understand complex traits. Nat Rev Genet 15, 34–48 (2014).

41. Wray, N. R., Wijmenga, C., Sullivan, P. F., Yang, J. & Visscher, P. M. Common Disease Is More Complex Than Implied by the Core Gene Omnigenic Model. Cell 173, 1573–1580 (2018).

42. Koivula, R. W. et al. Discovery of biomarkers for glycaemic deterioration before and after the onset of type 2 diabetes: descriptive characteristics of the epidemiological studies within the IMI DIRECT Consortium. Diabetologia 62, 1601–1615 (2019).

43. Delaneau, O. & Marchini, J. Integrating sequence and array data to create an improved 1000 Genomes Project haplotype reference panel. Nat Commun 5, 1–9 (2014).

44. Howie, B. N., Donnelly, P. & Marchini, J. A Flexible and Accurate Genotype Imputation Method for the Next Generation of Genome-Wide Association Studies. PLoS Genet 5, (2009).

45. Initial sequencing and analysis of the human genome. Nature 409, 860–921 (2001).

46. Marco-Sola, S., Sammeth, M., Guigó, R. & Ribeca, P. The GEM mapper: fast, accurate and versatile alignment by filtration. Nat Methods 9, 1185–1188 (2012).

47. Harrow, J. et al. GENCODE: the reference human genome annotation for The ENCODE Project. Genome Res. 22, 1760–1774 (2012).

48. Delaneau, O. et al. A complete tool set for molecular QTL discovery and analysis. Nat Commun 8, 1–7 (2017).

49. The GEUVADIS Consortium et al. Reproducibility of high-throughput mRNA and small RNA sequencing across laboratories. Nat Biotechnol 31, 1015–1022 (2013).

50. Fort, A. et al. MBV: a method to solve sample mislabeling and detect technical bias in large combined genotype and sequencing assay datasets. Bioinformatics 33, 1895–1897 (2017).

51. Li, Y. I. et al. LeafCutter: annotation-free quantification of RNA splicing. http://biorxiv.org/lookup/doi/10.1101/044107 (2016) doi:10.1101/044107.

52. Assarsson, E. et al. Homogenous 96-Plex PEA Immunoassay Exhibiting High Sensitivity, Specificity, and Excellent Scalability. PLOS ONE 9, e95192 (2014).

53. Ongen, H., Buil, A., Brown, A. A., Dermitzakis, E. T. & Delaneau, O. Fast and efficient QTL mapper for thousands of molecular phenotypes. Bioinformatics 32, 1479–1485 (2016).

54. Brown, A. A. LargeQvalue: A Program for Calculating FDR Estimates with Large Datasets | bioRxiv. https://www.biorxiv.org/content/10.1101/010074v2.

55. GTEx Consortium. Genetic effects on gene expression across human tissues. Nature 550, 204–213 (2017).

56. Bates, D., Mächler, M., Bolker, B. & Walker, S. Fitting Linear Mixed-Effects Models Using lme4. Journal of Statistical Software 67, 1–48 (2015).

57. Saha, A. & Battle, A. False positives in trans-eQTL and co-expression analyses arising from RNA-sequencing alignment errors. F1000Res 7, (2019).

58. Brown, A. A. et al. Predicting causal variants affecting expression by using whole-genome sequencing and RNA-seq from multiple human tissues. Nature Genetics 49, 1747–1751 (2017).

59. Scutari, M. Learning Bayesian Networks with the bnlearn R Package. arXiv:0908.3817 [stat] (2010).

60. Giambartolomei, C. et al. Bayesian Test for Colocalisation between Pairs of Genetic Association Studies Using Summary Statistics. PLoS Genet 10, e1004383 (2014).

61. Klimentidis, Y. C. et al. Phenotypic and Genetic Characterization of Lower LDL Cholesterol and Increased Type 2 Diabetes Risk in the UK Biobank. Diabetes 69, 2194–2205 (2020).

62. Hoffmann, T. J. et al. A large electronic-health-record-based genome-wide study of serum lipids. Nature Genetics 50, 401–413 (2018).

63. Shannon, P. et al. Cytoscape: A Software Environment for Integrated Models of Biomolecular Interaction Networks. Genome Res. 13, 2498–2504 (2003).

64. Ou, J. & Zhu, L. J. trackViewer: a Bioconductor package for interactive and integrative visualization of multi-omics data. Nature Methods 16, 453–454 (2019).

