## Supplementary Figures and Tables for "Genetic analysis of blood molecular phenotypes reveals regulatory networks affecting complex traits: a DIRECT study"

**Supplementary Tables ……………………………………………………………………..2**
**Supplementary Figures ………………………………………………………………….3**

| Phenotype | N. Samples | Male/Female | N. Phenotypes |
|---|---|---|---|
| **Genotypes** | 3,029 | 2142/887 | 9,472,419 |
| **Gene expression** | 3,029 | 2142/887 | 16,205 |
| **Exon expression** | 3,029 | 2142/887 | 140,198 |
| **Splicing phenotypes** | 3,029 | 2142/887 | 64,546 |
| **Targeted proteome** | 3,015 | 2134/881 | 373 |
| **Targeted metabolites** | 3,029 | 2142/887 | 116 |
| **Untargeted metabolites** | 2,998 | 2121/887 | 233 |

**Supplementary Table 1 |** After quality evaluation of all molecular phenotypes, the full dataset was restricted to individuals with complete RNAseq-genotypes data (N=3,029). Full break down of the number of participants for study with the different molecular phenotypes is included here. All analyses were performed with the maximum possible number of participants, which oscillates between 2,998 and 3,029.

| Phenotype class | Phenotypes | At least 1 association | More than 2 associations | QTLs |
|---|---|---|---|---|
| Cis-eQTLs (genes) | 16,205 | 15,305 (94.4%, pi1=0.968) | 12,824 | 59,972 |
| Cis-eQTLs (exons) | 140,198 (15,712) | 14,759 (93.9%, pi1 = 0.968) | 12,477 | 59,952 |
| Cis-sQTLs (splicing) | 64,546 | 3,609 (5.59%, pi1=0.655) | 298 | 3,978 |
| Cis-pQTLs (targeted Proteins) | 373 | 363 (97.31% , pi1=0.98) | 315 | 1,590 |
| Trans-eQTLs (genes) | 15,114 | 1,670 (11.04%, pi1=0.513) | 344 | 2,320 |
| Trans-pQTLs (targeted proteins) | 373 | 139 (37.2%, pi1=0.463) | 95 | 533 |
| metaboQTLs (targeted metabolites) | 116 | 69 (65.09%, pi1=0.813) | 33 | 102 |
| metaboQTLs (untargeted metabolites) | 233 | 103 (44.2%, pi1=0.535) | 96 | 199 |

**Supplementary Table 2 | QTL discovered in DIRECT**. Number of significant QTL identified for the different molecular phenotypes are indicated. From RNAseq data we derived gene expression quantifications, exon expression quantifications and splicing phenotypes. In addition, we used targeted proteins, targeted metabolites and untargeted metabolites. The analyses were done considering nearby SNPs (cis-QTLs) for expression and protein phenotypes with a genomics location, and considering SNPs acting in trans (more than 5Mb from TSS of the phenotypes). Metabolites, with no specific genomic location were assessed for associations with all the SNPs. The columns indicate the type of phenotypes (Phenotype class) and QTLs derived, the number of phenotypes (Phenotypes) tested in each analysis, the number of phenotypes with at least one significant association, the number of phenotypes with more that two significant associations after performing the conditional association analysis, and the final total number of associations (QTLs) per phenotype.

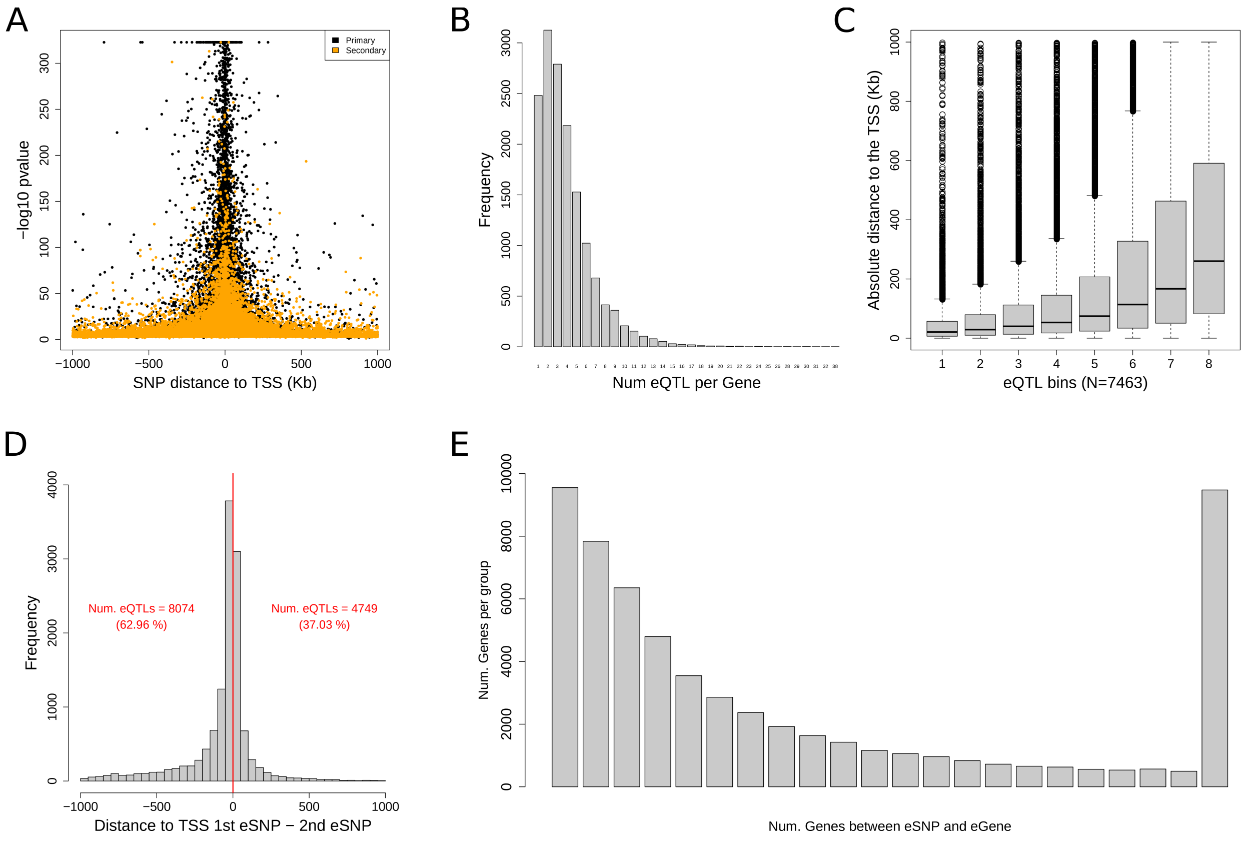


**Supplementary Figure 1 | Properties of cis-eQTLs. A)** Most eSNPs were located near the TSS of the genes. The plot shows the location of the lead eSNP per cis-eQTL (x-axis) and the -log10 of the pvalues for association of the 15,305 genes (94.4%) with at least one eQTL in black (n=15,305). Orange points identify any independent secondary cis-eQTL (n=44,667). **B)** Majority of genes have more than one cis-eQTLs (mean = 3). The barplot shows the number of cis-eQTLs per gene. Genes with two or more significant independent associations were 83.7% (12,824) of the genes with at least one cis-eQTL. **C)** Primary gene level cis-eQTLs, identify as those cis-eQTLs with the most significant association of the multiple independent signals, were significantly closer from the canonical TSS of the genes than secondary signals (Wilcox test = 7.3492e-173). The plot shows the absolute distance between the eSNPs and the TSS for all cis-eQTLs (y-axis), grouped in sets of 7,463 eSNPs with similar pvalue: group 1 had the largest pvalues, while group 8 had the smaller, but significant pvalues. **D)** The histogram show the distance between the 1$^{st}$ and the 2$^{nd}$ eSNPs for genes with two cis-eQTLs. Positive distance indicated the 1$^{st}$ and most significant cis-eQTLs (37.03% of cis-eQTLs) was further way form the TSS of the gene. **E)** Barplot showing the number of other genes which canonical TSS was located between the eSNP and the genes of all the independent cis-eQTLs. For 84.66% of the cis-eQTLs, we would identify at least one gene (as defined by Gencode v19) located between the variant and the gene of the cis-eQTL.

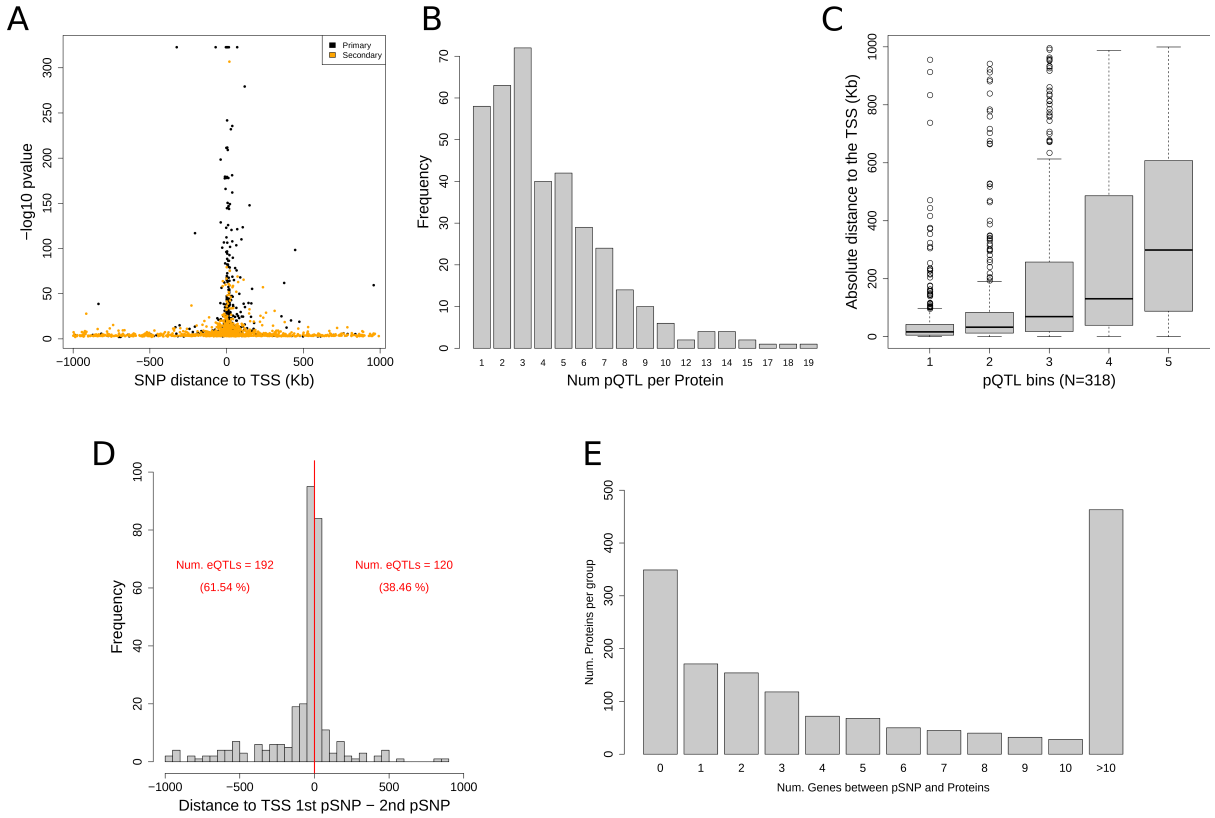


**Supplemental Figure 2 | Properties of cis-pQTLs are shared with cis-eQTLs. A)** Like cis-eQTLs, the location of the SNPs acting as cis-pQTLs (pSNPs) centred around the TSS of the coding gene for the proteins. The plot shows in black these locations for leads cis-pSNP (x-axis) and the -log10 pvalues for association of the cis-pQTL. Likewise, orange points identify any independent secondary cis-pQTL. **B)** The histogram shows the distribution of independent pQTLs per protein. Of the proteins with one pQTL, 86.7% of them had at least one secondary pQTLs (84.4% of all proteins). C) Similarly to cis-eQTLs, the stronger cis-pQTLs were significantly closer from the canonical TSS of the gene associated to the protein than secondary signals (Wilcoxon test = 9.54e-25). **D)** Also proteins with at least two cis-pQTLs presented secondary associations closer to the TSS than the primary associations. The proportion were of 38.46% cis-pQTLs (120) were the secondary pQTLs was closer to the TSS than the primary cis-pQTL. **E)** Barplot showing the number of other genes which canonical TSS was located between the pSNP and the genes of all the independent cis-pQTLs. For 78.05% of the cis-pQTLs, we would identify at least one coding gene for the proteins (as defined by Gencode v19) located between the variant and the gene of the cis-pQTL.

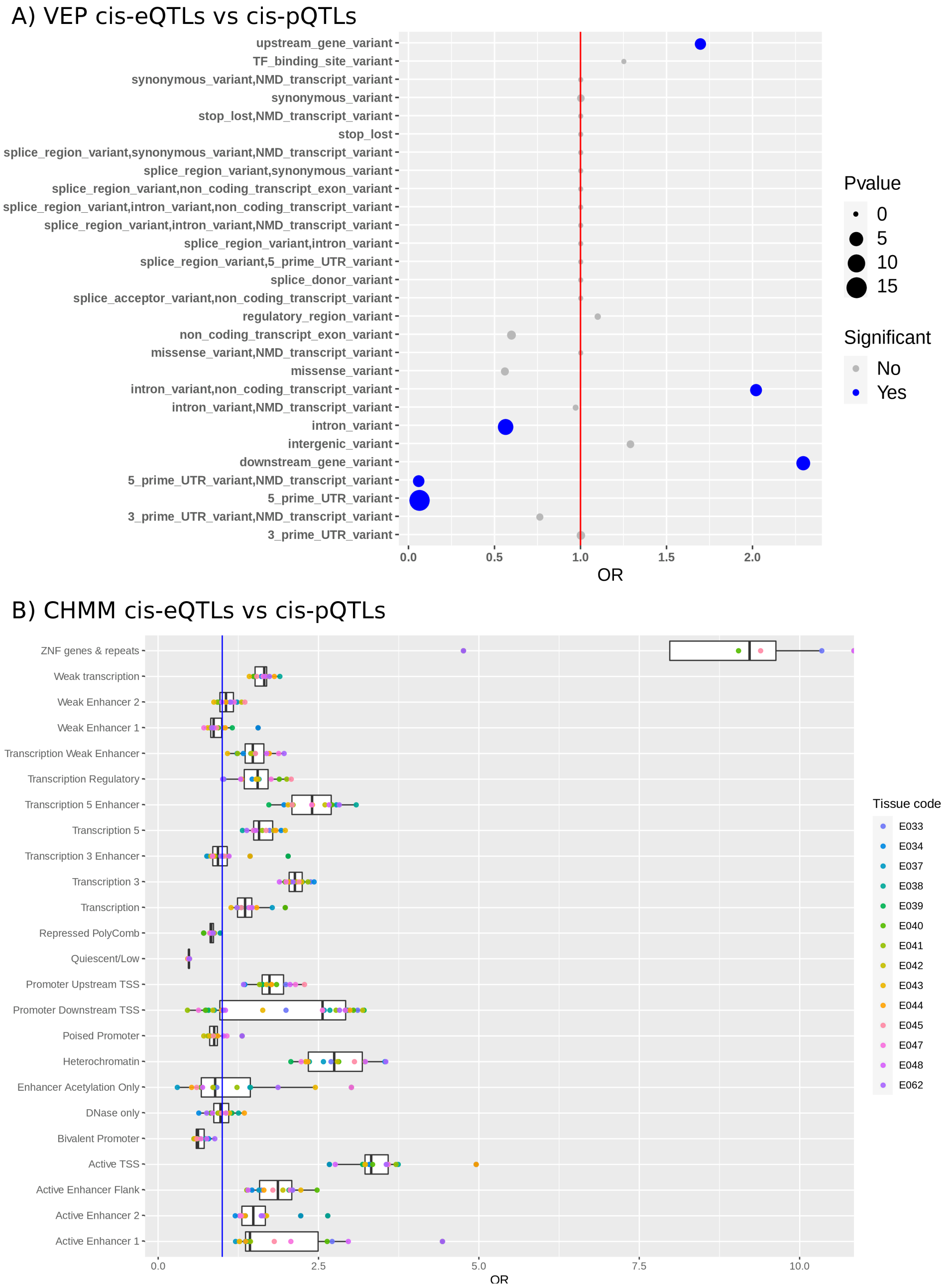


**Supplemental Figure 3 | Functional annotation and enrichment analysis of cis-eSNPs vs. cis-pSNPs. A)** Functional enrichment analysis of cis-eSNP vs. cis-pSNPs using VEP. Positive OR indicates enrichment in cis-eSNPs. **B)** Functional enrichment analysis using ChromHMM experiments. Cell types included were: E062 (Primary mononuclear cells from peripheral blood), E034 (Primary T cells from peripheral blood), E045 (Primary T cells effector/memory enriched from peripheral blood), E033 (Primary T cells from cord blood), E044 (Primary T regulatory cells from peripheral blood), E043 (Primary T helper cells from peripheral blood), E039 (Primary T helper naive cells from peripheral blood), E041 (Primary T helper cells PMA-I stimulated), E042 (Primary T helper 17 cells PMA-I stimulated), E040 (Primary T helper memory cells from peripheral blood), E037 (Primary T helper memory cells from peripheral blood), E048 (Primary T CD8+ memory cells from peripheral blood), E038 (Primary T helper naive cells from peripheral blood), and E047 (Primary T CD8+ naive cells from peripheral blood).

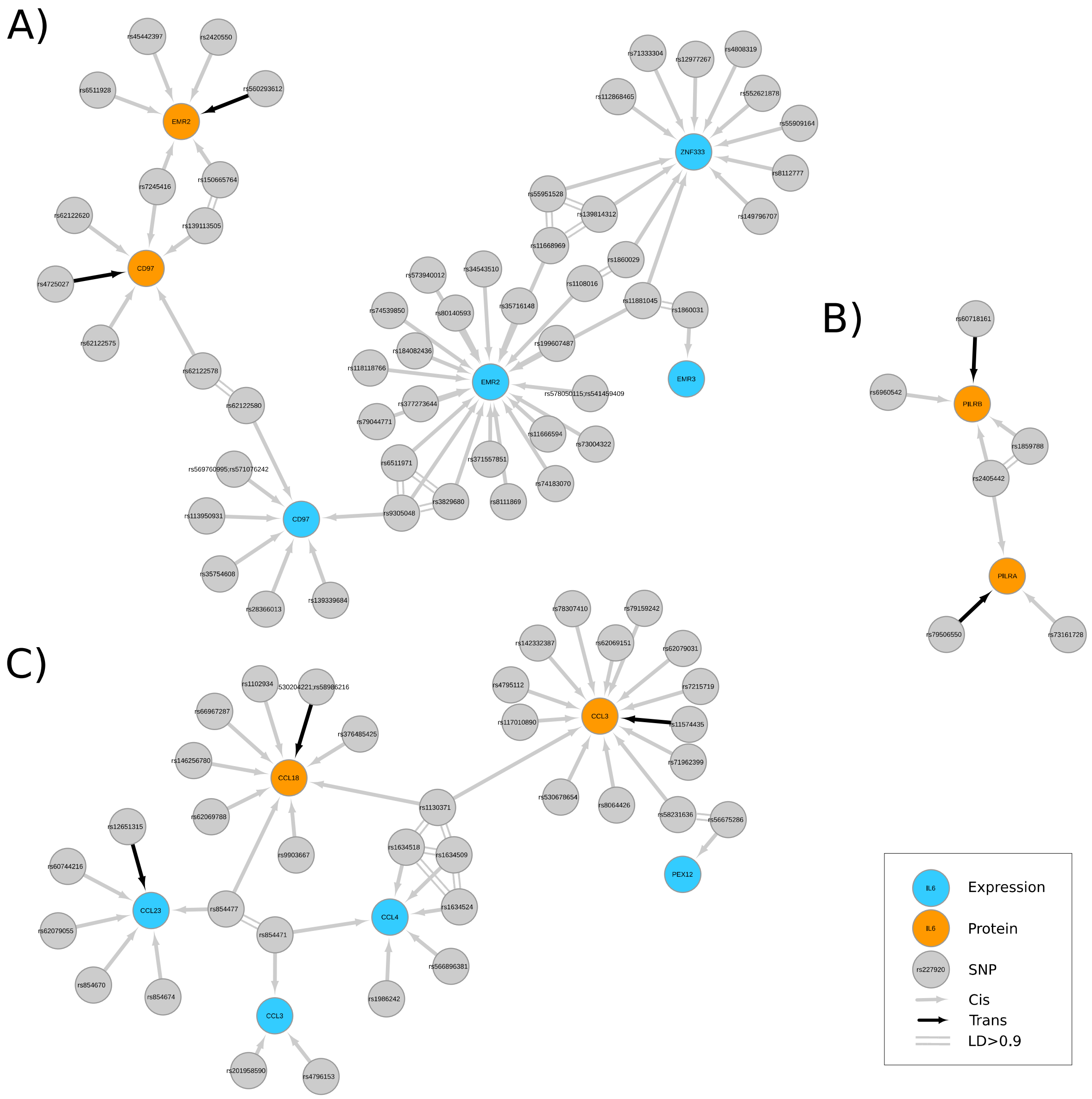


**Supplementary Figure 4 | Pleiotropy in cis-pQTLs.** We highlight here 3 networks showing cis-pSNPs associated to more than one protein. The figure shows all independent networks associated to the SNPs. Proteins are represented as orange nodes, SNPs as grey nodes and gene expression as blue nodes. Grey arrows indicate significant cis QTLs for proteins or genes, black arrows identify trans-QTLs. **A)** Network for CD97 and EMR2 on chromosome 19 with a pSNP (rs7245416) shared that deceased their abundances. The local cis-network also connects with the expression of CD97 and EMR2. **B)** Network of cis-pQTLs for PILRB and PILRA in chromosome 7 sharing one cis-pQTL (rs2405442). **C)** Network of QTLs for CCL18 and CCL3 on chromosome 17 with one pSNP (rs1130371) in common increasing the abundance of both proteins.

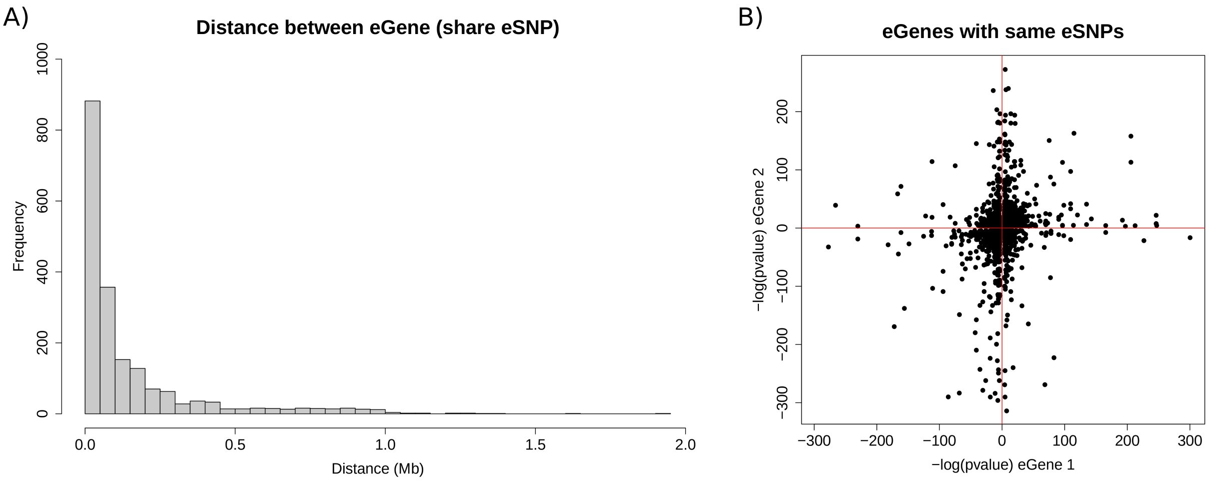


**Supplementary Figure 5 | Properties of eGenes sharing eSNPs. A**) Genes that were regulated by the same eSNP were often close to each other. The histogram shows the abundance of genes sharing the lead cis-eSNPs ordered by the distance between the TSS of the pair of genes in Mb. The mean distance between the 1,924 pairs where SNPs or pairs of SNPs in high LD (R2>0.9) associated with two different genes was 0.14 Mb. **B**) Shared cis-eSNP effects had more often opposite direction of effects when they were further away from each other. The plot shows the –log10 pvalue adjusted by the sign of the slope (direction of effect of the alternative allele) for gene 1 (x-axis) vs gene 2 (y-axis). Of the pairs of genes, 583 cis-eQTLs (30.30%) showed opposite direction of effect despite the same eSNP being involved and were more likely to be further away that those with similar direction of effect (Wilcoxon test P = 7.16e-15). Genes that overlap an exon in the same or in opposite strand were removed for this analysis.

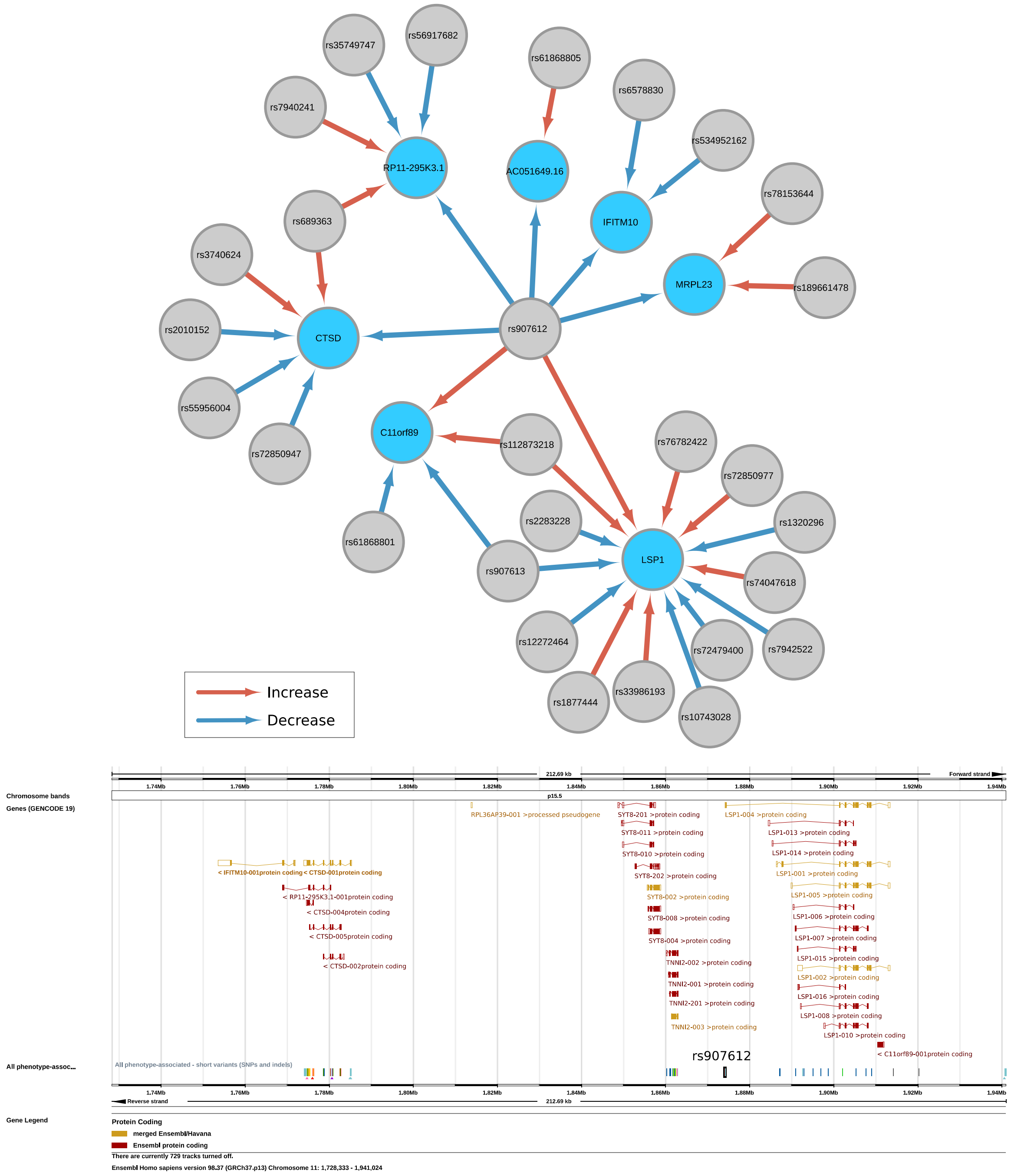


**Supplementary Figure 6 | cis-eSNPs regulate the expression of different genes with opposite direction of effect.** An example of shared cis-eQTLs (rs907612) involving multiple genes were the same allele had a different direction of effects in chromosome 11. Examples of the effect allele T (rs907612-T) increasing expression are indicated with red arrows: *LSP1* and *C11orf89.* Examples were rs907612-T degreased expression (blue arrows) included *IFITM10*, *CTSD*, *MRPL12*, *RP11-295K3.1*, *AC051649.16.* The genomic region, indicated at the bottom, shows the location of rs907612 (black) and other SNPS.

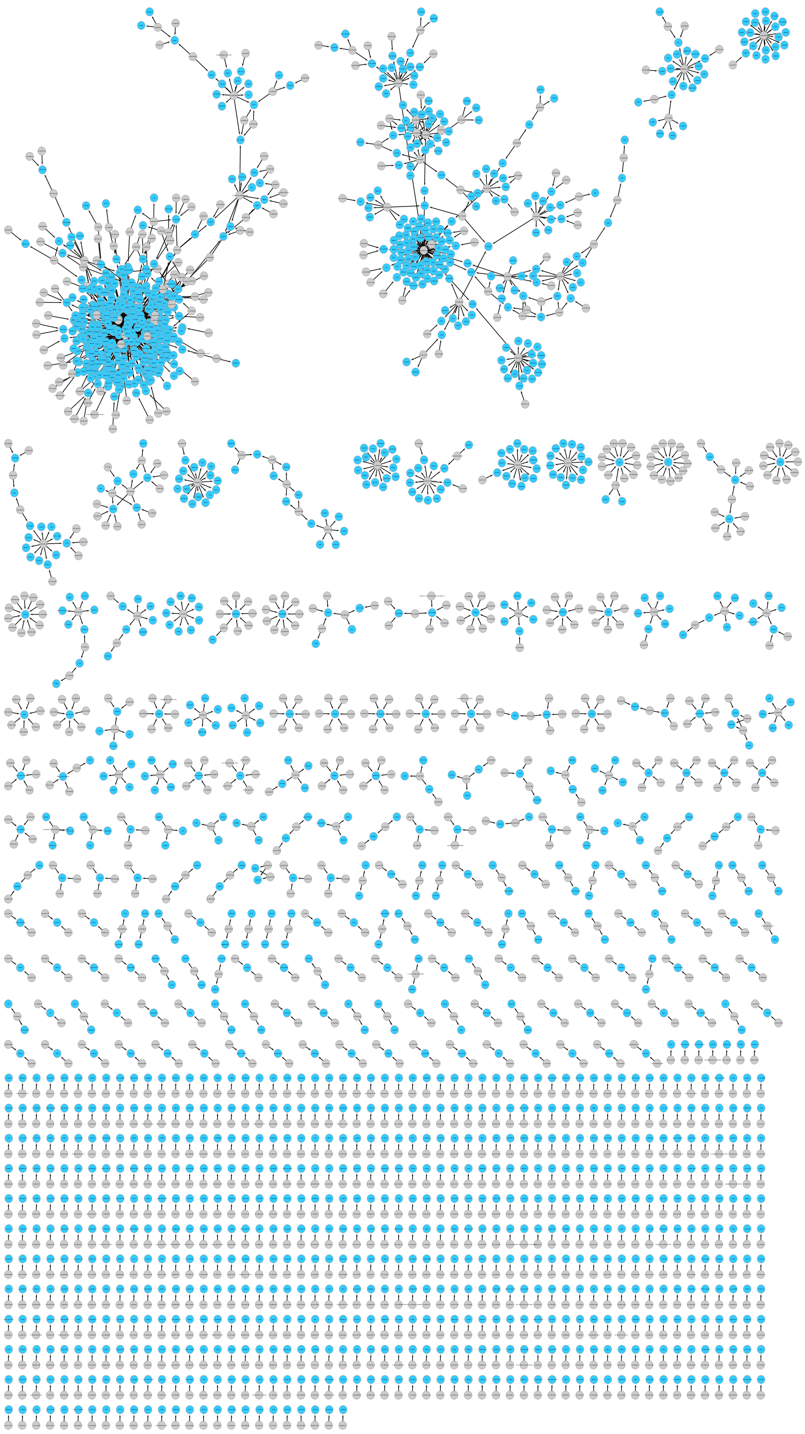

**Supplementary Figure 7 | Visualization of significant trans-eQTLs.** Genes are represented as blue nodes while trans-eSNPs (lead SNPs associated to the expression of a gene in trans) are grey nodes. We identified 2,320 independent trans-eQTLs (Supplementary Data 6), involving 1,670 genes and 1,351 SNPs (from the 15,115 genes tested). Of the genes with a trans-eQTL (trans-eGenes), 345 (20.65%) had more than one trans-eSNP independently regulating their expression (mean = 1.39), with a maximum of 13 distal eSNP effects for *F7*.

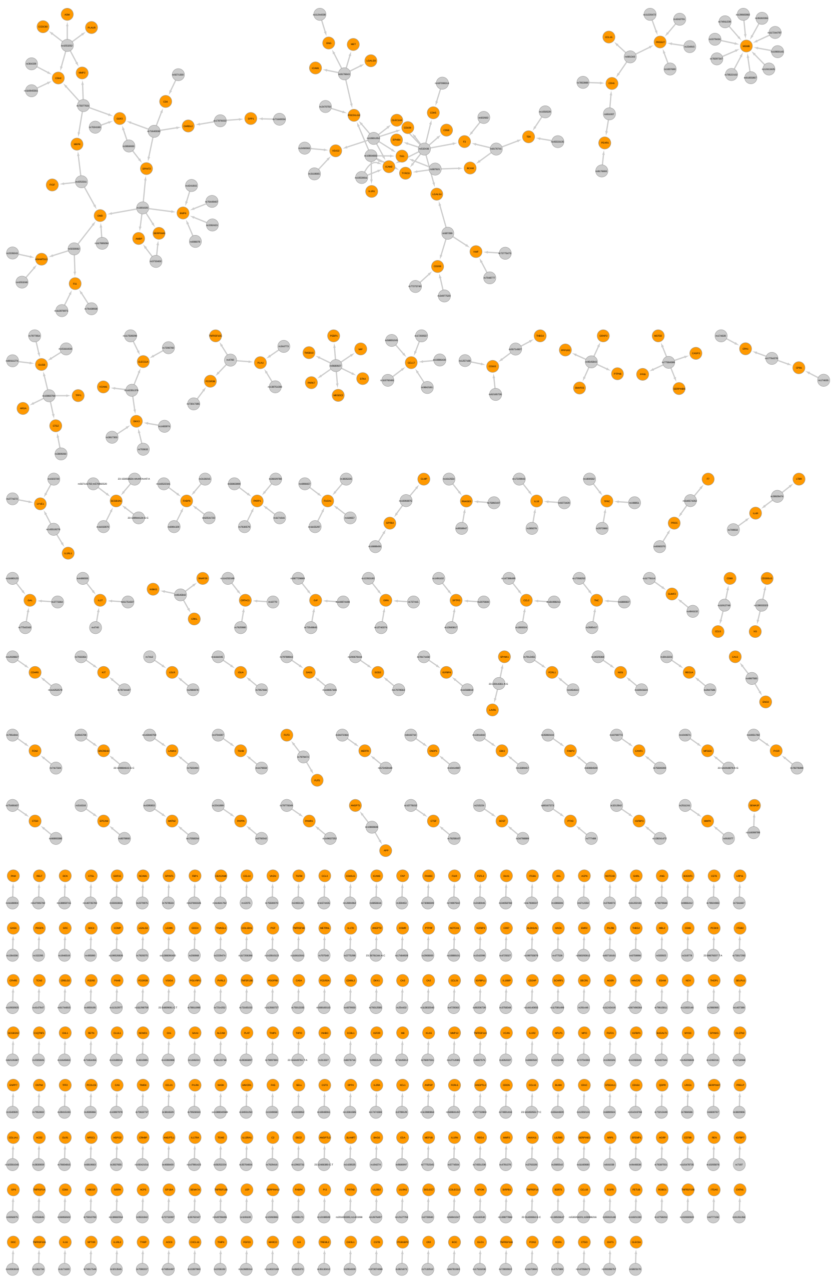

**Supplementary Figure 8 | Visualization of significant trans-pQTLs.** This visualization of trans-pQTL present proteins as orange nodes and trans-pSNPs (lead SNPs associated to the proteins in trans) as grey nodes. Our analysis identified 533 independent trans-pQTLs 193 proteins. We would observe a similar abundance of allelic heterogeneity than for trans expression associations. The number of proteins being regulated by more than one trans-pSNP was slightly larger than observed for expression.

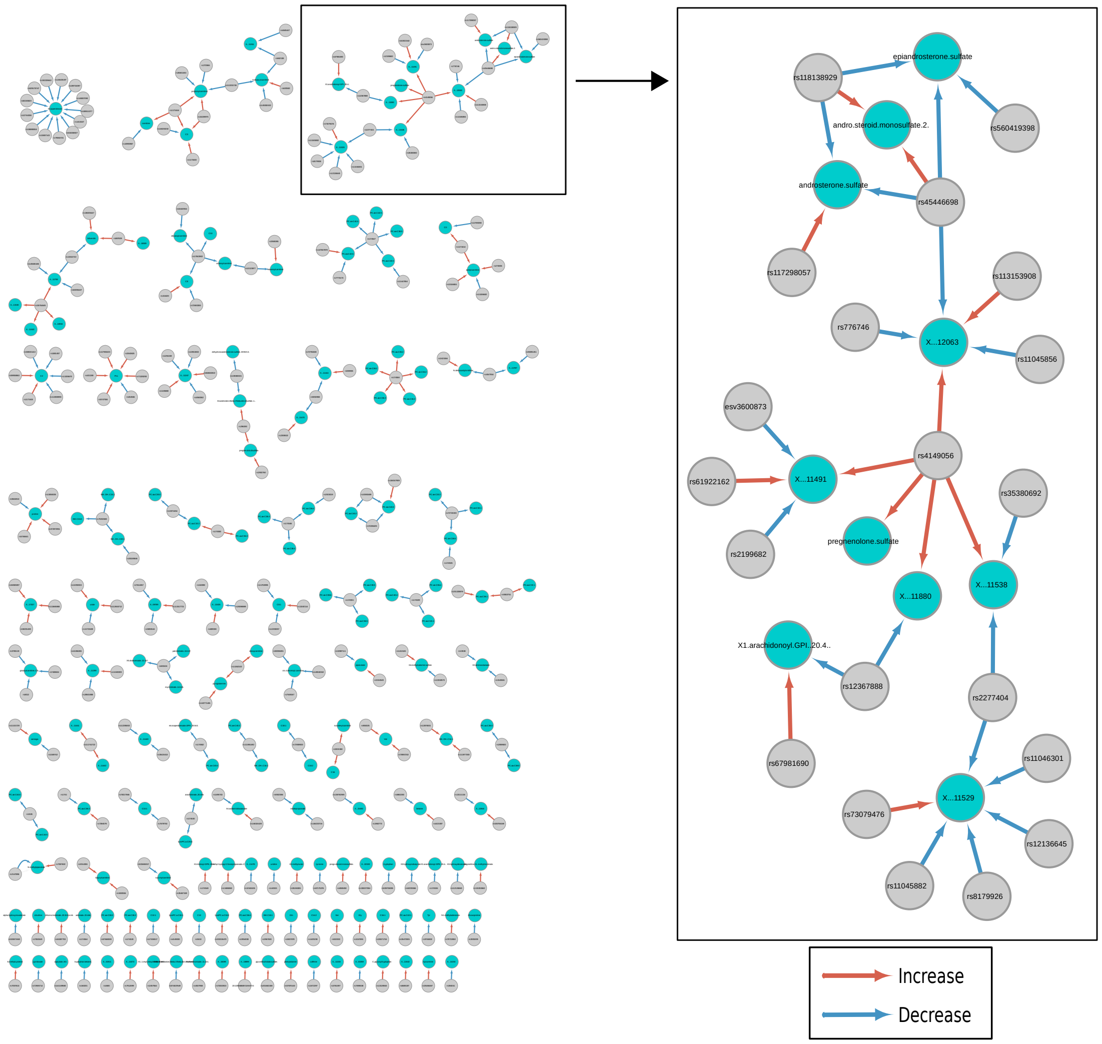


**Supplementary Figure 9 | Visualization of significant metabo-QTLs. A)** A genome-wide scan of metabo-QTLs identified 301 independent associations for 172 targeted and untargeted metabolites (37.47%). Of the associated metabolites 68 have more than one metabo-QTLs (39.5%, mean=1.75), with up to 13 SNPs for 2-piperidinone. Likewise with other molecular phenotypes, we identify variants (n=42) associated to two or more metabolites (mean=1.27). **B)** Highlight of one of the networks derived from metabo-QTLs with a SNP associated with up to 5 metabolites (rs174547).

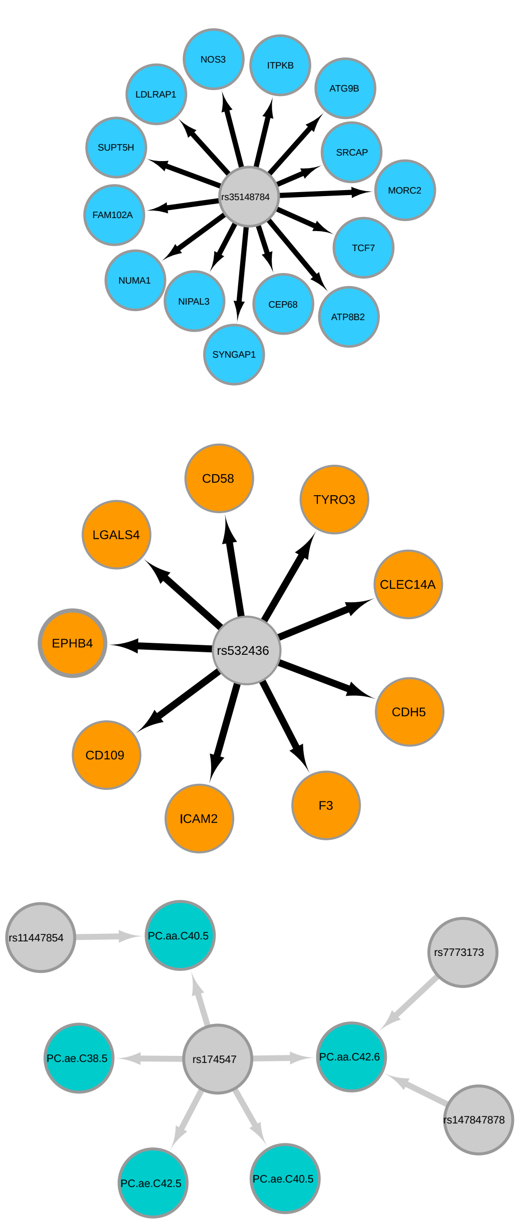


**Supplemental Figure 10 | Trans-QTLs associate with multiple phenotypes.** Examples of trans-SNPs associated to multiple phenotypes. **A and B)** Trans-eQTLs association for rs35148784 (grey node) with the expression of 14 genes (blue nodes), and associations in trans for rs4065321 with the expression of 16 genes. **C and D)** Trans-pQTL associations for rs532436 with 9 proteins (orange nodes) and rs9680927 and 6 proteins. **E and F)** Metabo-QTL associations for rs174547 and rs174584.

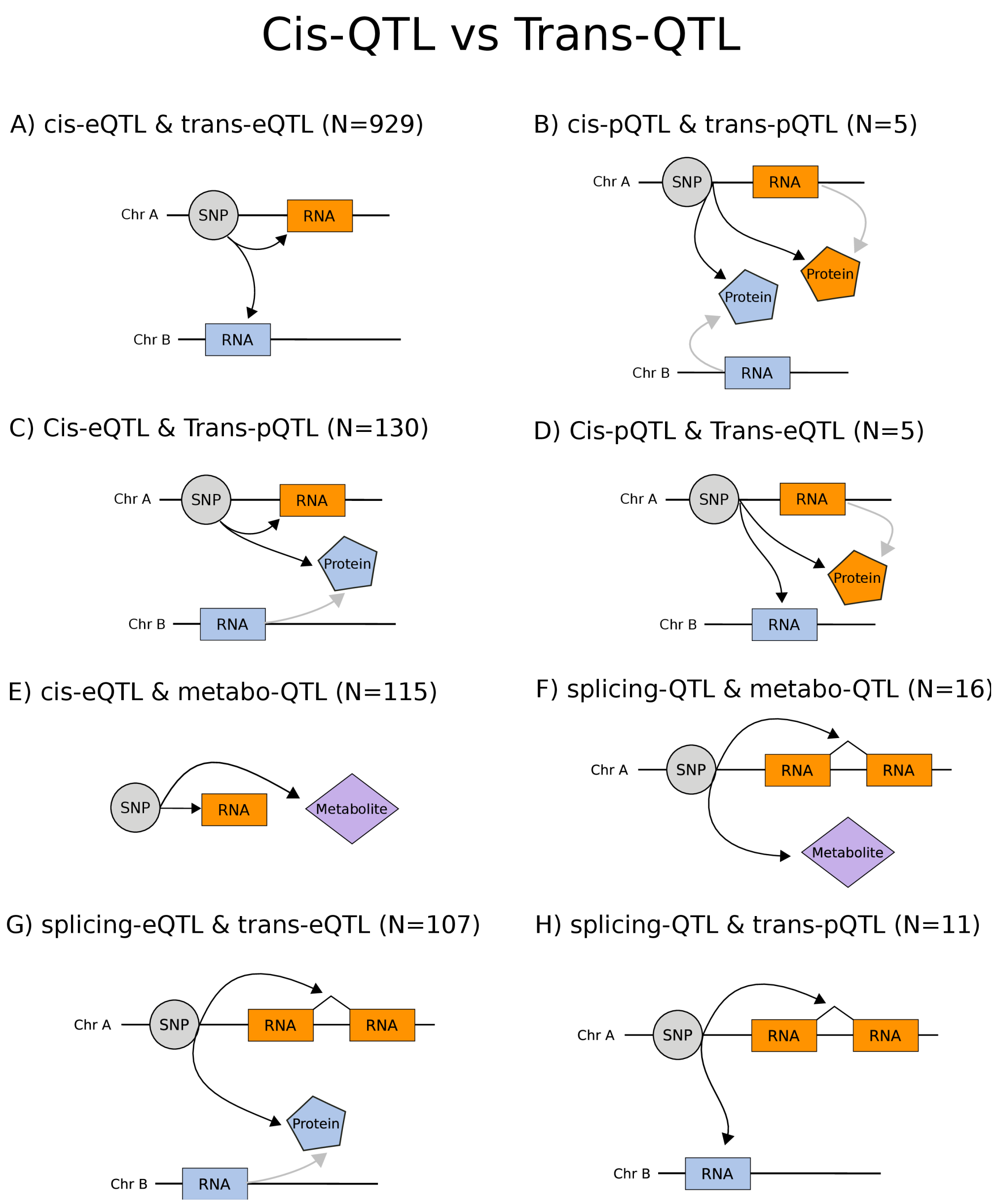


**Supplementary Figure 11 | cis-SNPs acting also as trans-SNPs.** From the significant QTL associations, we reported how many SNPs acted both locally (in cis) and distantly (in trans). We considered also pairs of SNPs in high LD ($R^2$>0.9), and metabo-QTL considered as trans-QTLs. Each of the diagrams show the SNP-phenotype pairs with multiple cis- and trans-QTLs. **A)** The number of trans-eQTLs that shared the same SNPs with cis-eQTLs were 262, including SNPs in high LD we identified 929 cases. **B)** Cis-pQTL that were also trans-pQTLs were 5, although we found no case were the exact same SNP acted both in cis and trans. **C)** SNPs acting on local gene expression and distal protein levels were 130 (26 reported same lead SNP). **D)** We identified 5 trans-eQTLs that were also cis-pQTLs, including rs34097845, a SNP in chromosome 17 associated locally with the MPO protein and acting distally to regulate the expression of both *HSPA5* (chromosome 9) and *HSP90B1* (chromosome 12) (Supplementary Figure 13A). **E)** Cis-eQTLs that were also acting as metabo-QTL were 115. **F)** splicing-QTLs shared associations with 16 metabo-QTLs; **G)** 107 associations with trans-eQTLs, and **H)** 11 with trans-pQTLs.

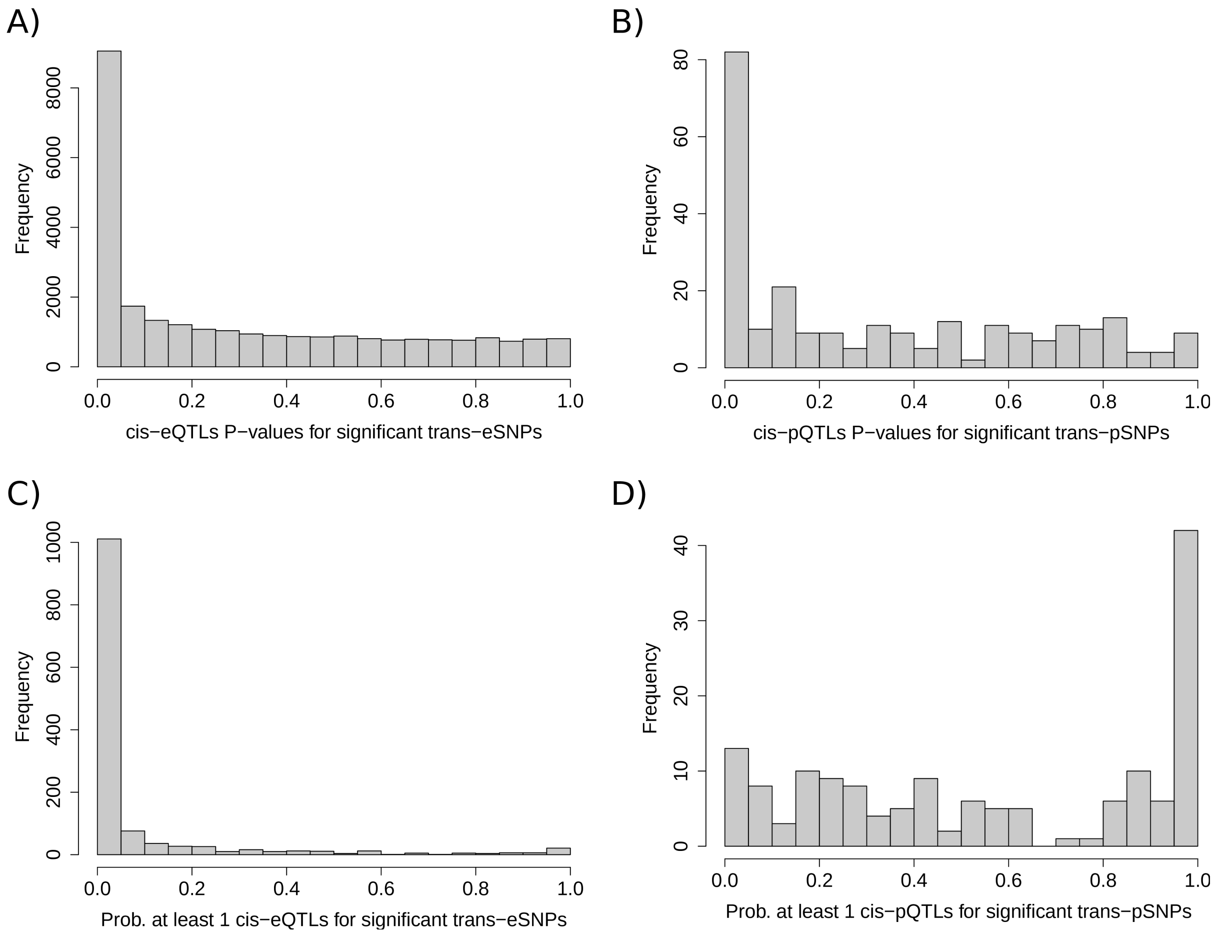


**Supplemental Figure 12 | Enrichment of cis-QTL in trans-SNPs. A)** Distribution of all cis-eQTL associations p-values from gene-SNPs pairs of significant trans-eSNPs. The $\pi_1$ enrichment using all pvalues (n=26,956) was of 41.42% for trans-eSNPs acting as cis-eQTLs. **B)** Distribution using all cis-pQTL Pvalues (n=253) from protein-SNPs pairs of significant trans-pSNPs. The $\pi_1$ enrichment was in this case of 41.68% for trans-pSNPs acting as cis-pQTLs. **C)** To account for multiple cis-QTL associations, we calculated the probability of trans-eSNPs being a cis-eQTL for at least one gene. The graph shows the Pvalue distribution (n=2,600). The $\pi_1$ enrichment identify 77.34% trans-eSNPs also acting as cis-SNP for at least one gene. **C)** Similarly to eQTLs, we plot the Pvalue distribution for the probability of trans-pSNPs being a cis-pQTL for at least one protein (n=153). The $\pi_1$ enrichment was of 0%, identify no trans-pSNPs acting also as cis-pQTLs.

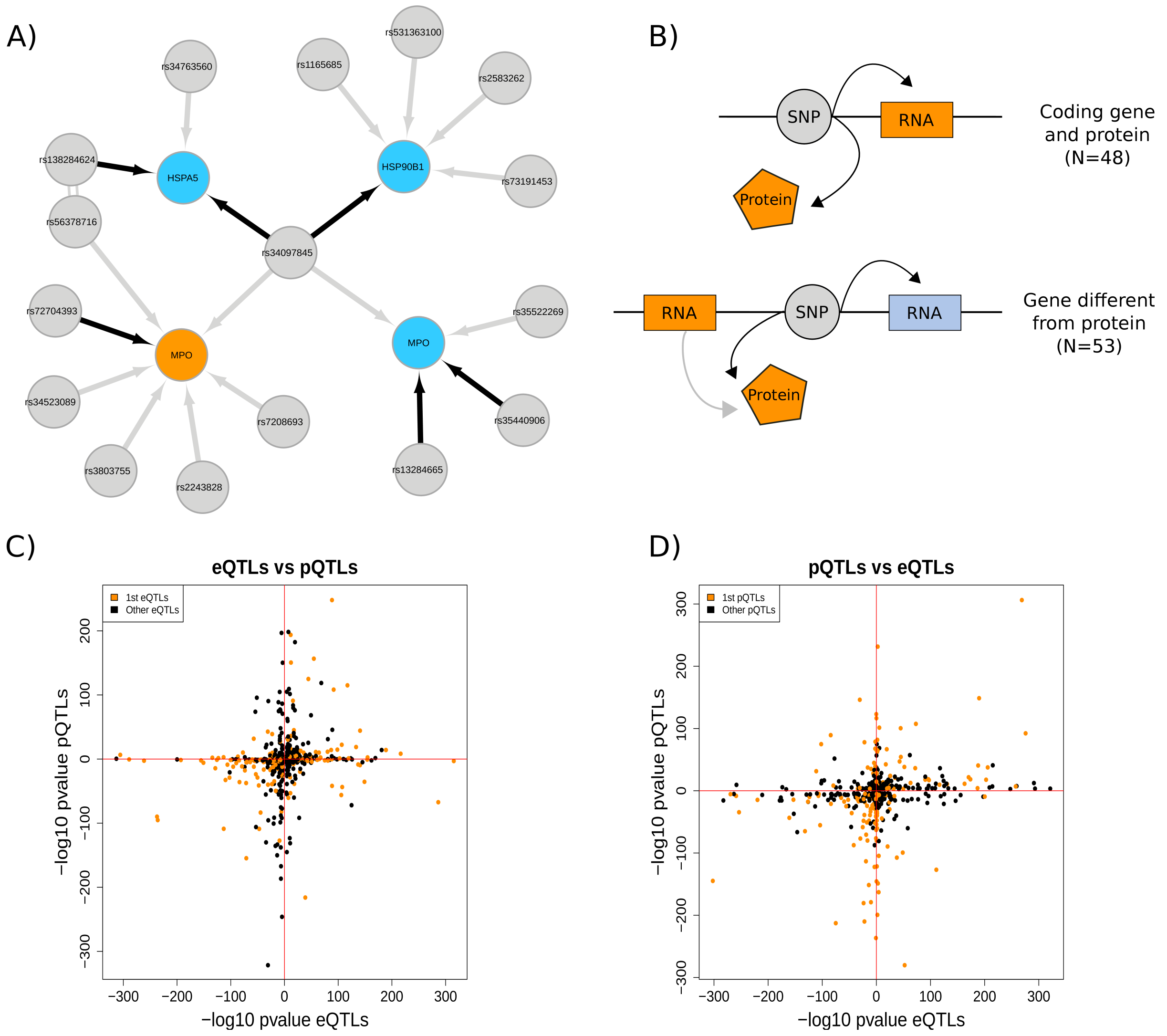


**Supplementary Figure 13 | Shared cis genetic regulation between genes expression and proteins. A)** rs34097845 was associated with both the expression of *MPO* and its protein (MPO) with a consistent direction of effect. The same SNPs is one of the 3 detected cases of a SNP acting as a cis-pQTL and a trans-eQTL. Proteins are presented as orange nodes, gene expression as blue nodes and SNPs as grey nodes. Cis associations are indicated by grey arrows, and trans in black. Double line indicate the SNPs were in high LD ($R^2$>0.9) **B)** We identified 101 trios of expression-SNP-proteins, of which 48 involved a protein and its coding gene, while 53 involved the expression of a nearby gene different that the coding gene for the protein. **C)** We evaluated the distributions of the P values for significant cis-eQTL, by extracting the Pvalue of cis-pQTLs for the same SNP-gene pair. Most cases were consistent in their direction of effect, with a few cases showing opposite direction of effect for the two phenotypes. **D)** We also evaluated the distributions of the P values for significant cis-pQTL, by extracting the pvalue of cis-eQTLs for the same SNP-gene pair observing the same trend.

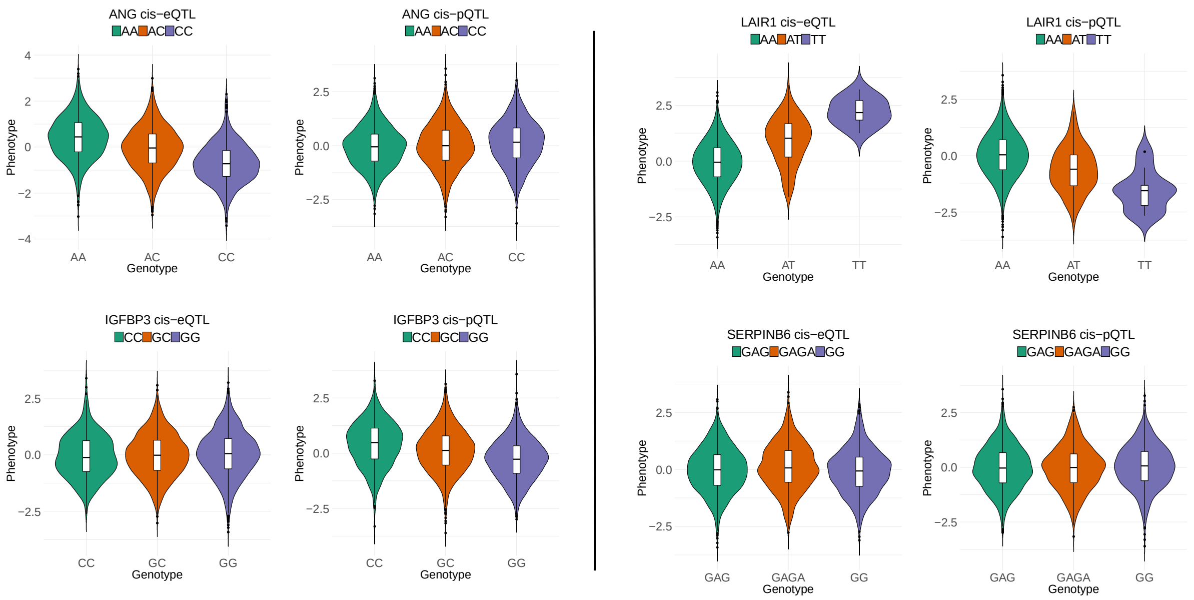


**Supplementary Figure 14 | Example of cis-SNPs regulating gene expression and proteins with opposite direction of effect. A)** The abundance of ANG and the expression of its coding gene *ANG* were significantly associated with rs1010461. For expression rs1010461-C decreased the expression of the gene, while for proteins the same allele increased their abundance. **B)** cis-QTLs for rs118056835 and the expression of *LAIR1* and ita protein. **C)** cis-QTLs for rs2854746 and the expression of *IGFBP3* and its protein. **D)** cis-QTLs for rs11335500 and the expression of *SERPINB6* and its protein.

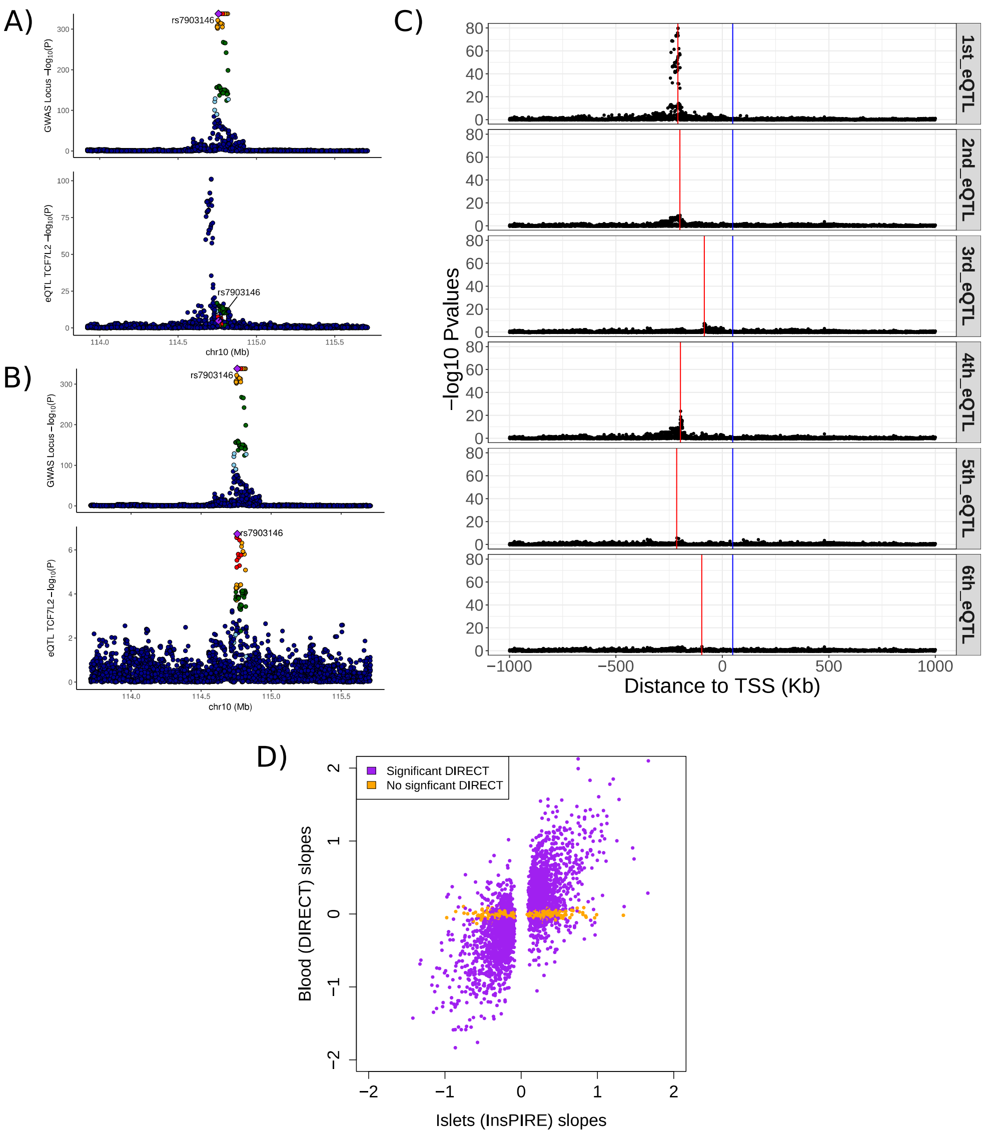


**Supplementary Figure 15 | cis-eQTLs for *TCF7L2*. A)** Comparison of associations between SNPs around the TSS of *TCF7L2* with T2D (top, Mahajan, et al. 2018) and its expression in DIRECT whole blood data (bottom). The lead GWAS variants (rs7903146) does not associate with the expression of *TCF7L2* in blood. **B)** Comparison of associations between SNPs around the TSS of *TCF7L2* with T2D (top) and its expression in pancreatic islets expression form the InsPIRE study (bottom, Viñuela et al, 2020). The lead GWAS variants (rs7903146) was significantly associated with the expression of *TCF7L2* in pancreatic islets. **C)** Regional plot for all the independent cis-eQTLs for *TCF7L2* in DIRECT blood. A total of 6 Independent cis-eQTLs were identified, indicated by red lines. The blue line identify the location of the T2D related GWAS variant (rs7903146). **D)** Comparative of cis-eQTLs from pancreatic islets (InsPIRE) and whole blood (DIRECT). From the significant cis-eQTLs in islets, we selected the -log10 pvalue from the same SNP-gene pair. A total of 486 genes were not significant in blood (Pvalue > 0.035, orange color) but significant in pancreatic islets (n=420). Of eQTLs significant in both studies (N = 2,691, Pvalue < 1e-05), 294 had opposite direction of effect.

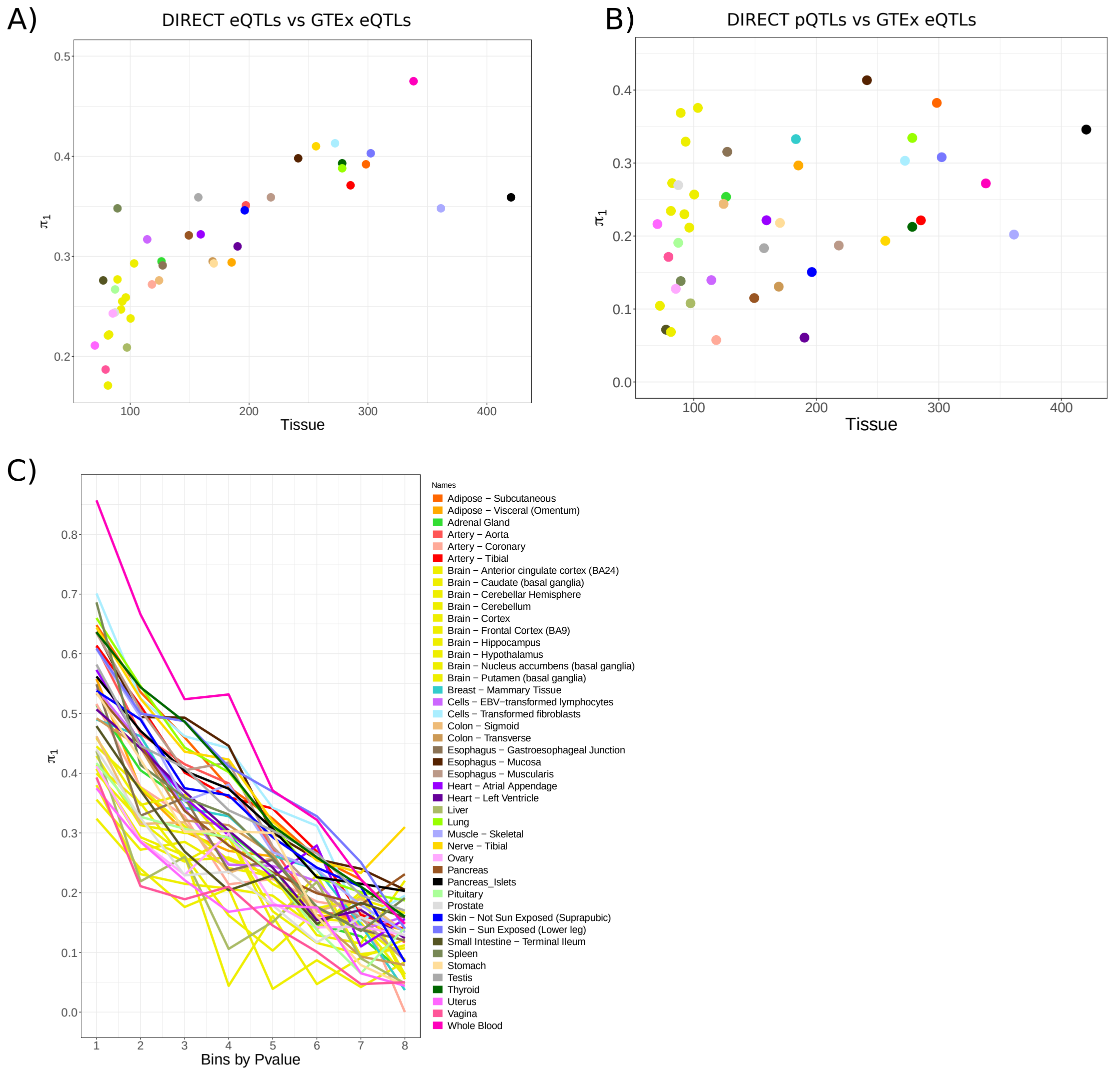


**Supplementary Figure 16 | GTEx comparison. A)** From the significant cis-eQTLs from DIRECT (n=3,029), we extracted the Pvalues for the same SNP-gene pairs from the GTEx and InsPIRE studies and plotted the $\pi_1$ enrichment values (y-axis) vs. the number of samples available per tissue. While Figure 2A shows how DIRECT identified most of the cis-eQTLs discovered in other studies, this figure shows how a large study is able to identify cis-eQTLs likely to be tissue specific and therefore not detected in other tissues with smaller sample size. **B)** From significant cis-pQTLs, we extracted Pvalues for the same SNP-gene pairs from the cis-eQTLs studies in other studies, plotting $\pi_1$ values (y-axis) vs. the number of samples available per tissue. We detected from 4.1% (heart, left ventricle) to 43.15% (adipose subcutaneous) of cis-pQTLs in blood shared genetic regulation with cis-eQTLs in other tissues. Here blood is no longer the tissue with the higher $\pi_1$, suggesting some of the proteins may have their origin in other tissues. **C)** From figure 2B, we order the independent cis-eQTLs Pvalues per gene from DIRECT by significance (bin1 most significant eQTLs) and reported $\pi_1$. The higher degree of shared associations across tissues were observed for the strongest cis-eQTLs and whole blood.

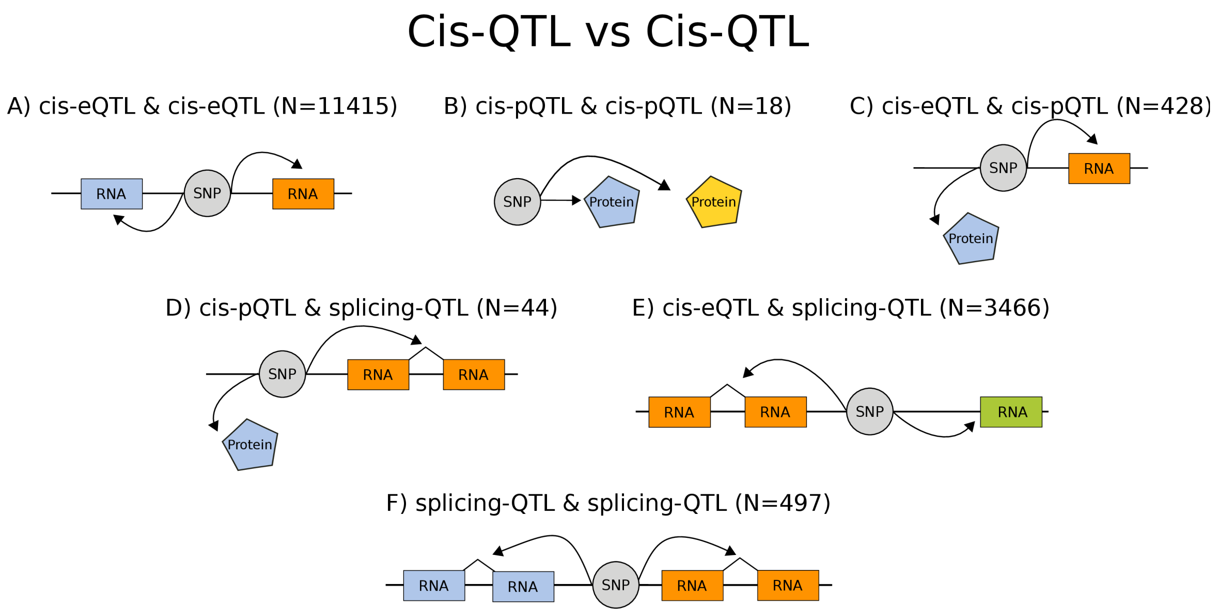


**Supplementary Figure 17 | Trios of cis-QTLs used to build a network of genetics perturbations across molecular phenotypes.** There were 6 possible combinations of cis-QTLs across expression, proteins and splicing phenotypes.

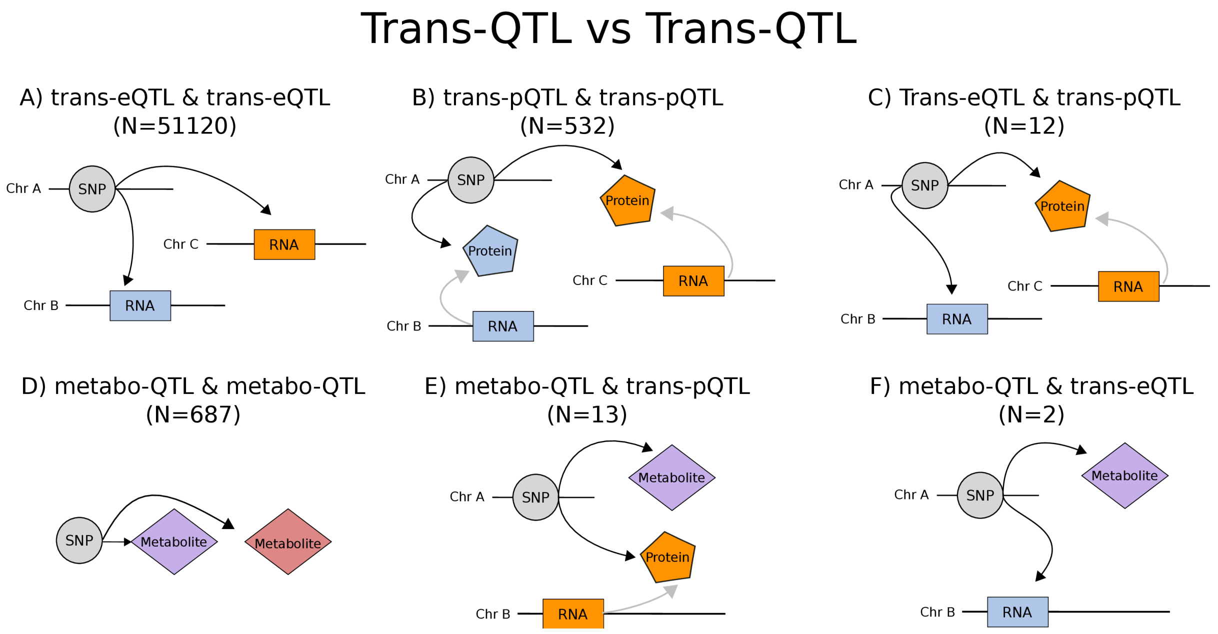


**Supplementary Figure 18 | Trios of trans-QTLs used to build a network of genetics perturbations across molecular phenotypes.** There were 6 possible combinations of trans-QTLs across, considering metabolites-QTLs as trans-QTLs, since they do not have an assigned genomic position.

### Cis-QTLs vs Cis-QTLs

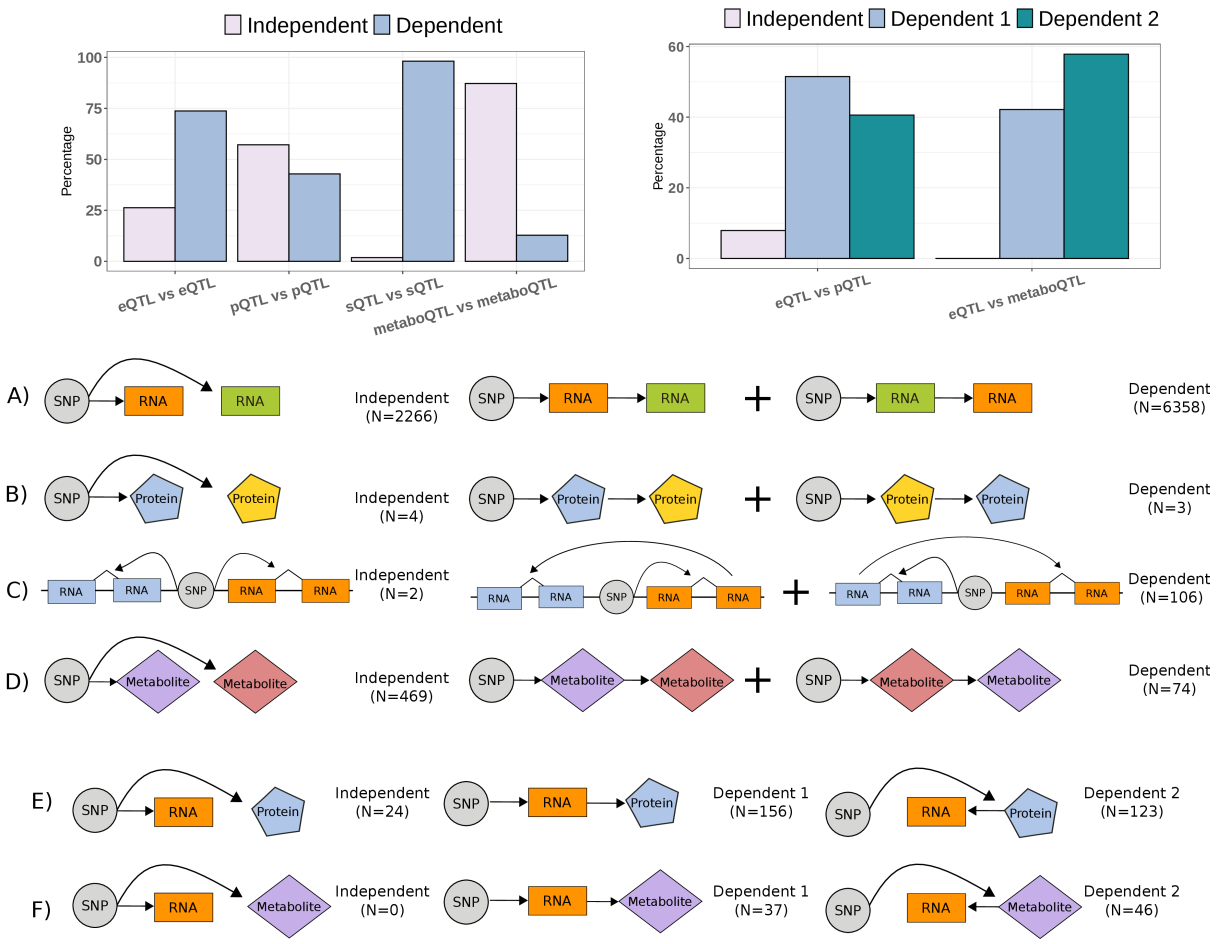


**Supplementary Figure 19 | Causal models for shared cis-QTLs.** Top barplots summarize the numbers indicated in diagrams A to F. These show cartoon representations of the models identified and tested for causality in cis. Colour code for the different models can be interpreted with the cartoons bellow. **A)** cis-eQTLs associated models, were the independent models evaluates unrelated effect of the SNPs in the expression of two genes and dependent models evaluates the dependent effect of the SNP on one gene mediated by the other gene. **B)** Models for cis-pQTLs. **C)** models for splicing-QTLs. **(D)** Models for metabolite-QTLs. **E)** Models for shared cis-eQTLs and cis-pQTLs. Here the dependent models distinguish between the effect of the SNP on proteins mediated by gene expression regulation (Dependent 1), and the effect of the SNP on expression mediated by proteins (Dependent 2). **F)** Models for shared cis-eQTLs and metabo-QTLs. Dependent models were for the effect of the SNP on metabolites mediated by gene expression regulation (Dependent 1), and the effect of the SNP on expression mediated by metabolites (Dependent 2).

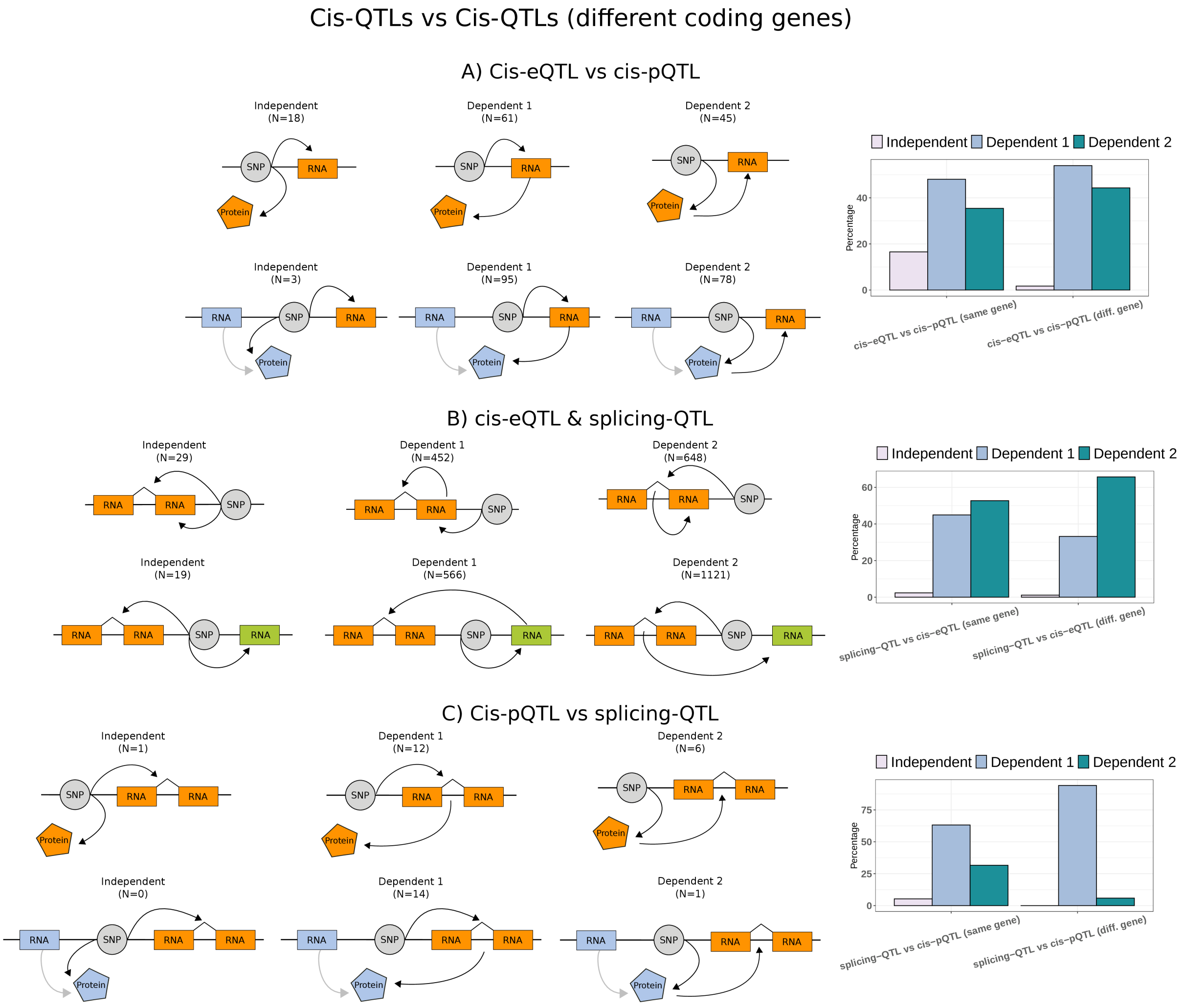


**Supplementary Figure 20 | Causal models for shared cis-QTLs (continuation).** Right barplots summarize the numbers indicated in diagrams A to C. These show cartoon representations of the models identified and tested for causality presented in Supplementary Figure 17 and separating cases were the SNP affected two phenotypes derived form the same gene or different gene. Colour code for the different models can be interpreted with the cartoons bellow. **A)** Models evaluating shared associations between expression and proteins. Top models indicated cases were the SNP affected protein levels and the expression of the coding gene. Here the dependent models distinguish between the effect of the SNP on proteins mediated by gene expression regulation (Dependent 1), and the effect of the SNP on expression mediated by proteins (Dependent 2). **B)** Models evaluating shared associations between expression and splicing. Top models indicated cases were the SNP affected both expression and splicing in the same gene. The dependent models distinguish between the effect of the SNP on gene splicing mediated by gene expression abundance regulation (Dependent 1), and the effect of the SNP on expression abundance mediated by a change in splicing (Dependent 2). **C)** Models evaluating shared associations between proteins and splicing. Top models indicated cases were the SNP affected both protein levels and splicing in the protein coding gene. The dependent models distinguish between the effect of the SNP on gene splicing mediated by protein abundance regulation (Dependent 1), and the effect of the SNP on proteins mediated by a change in splicing (Dependent 2).

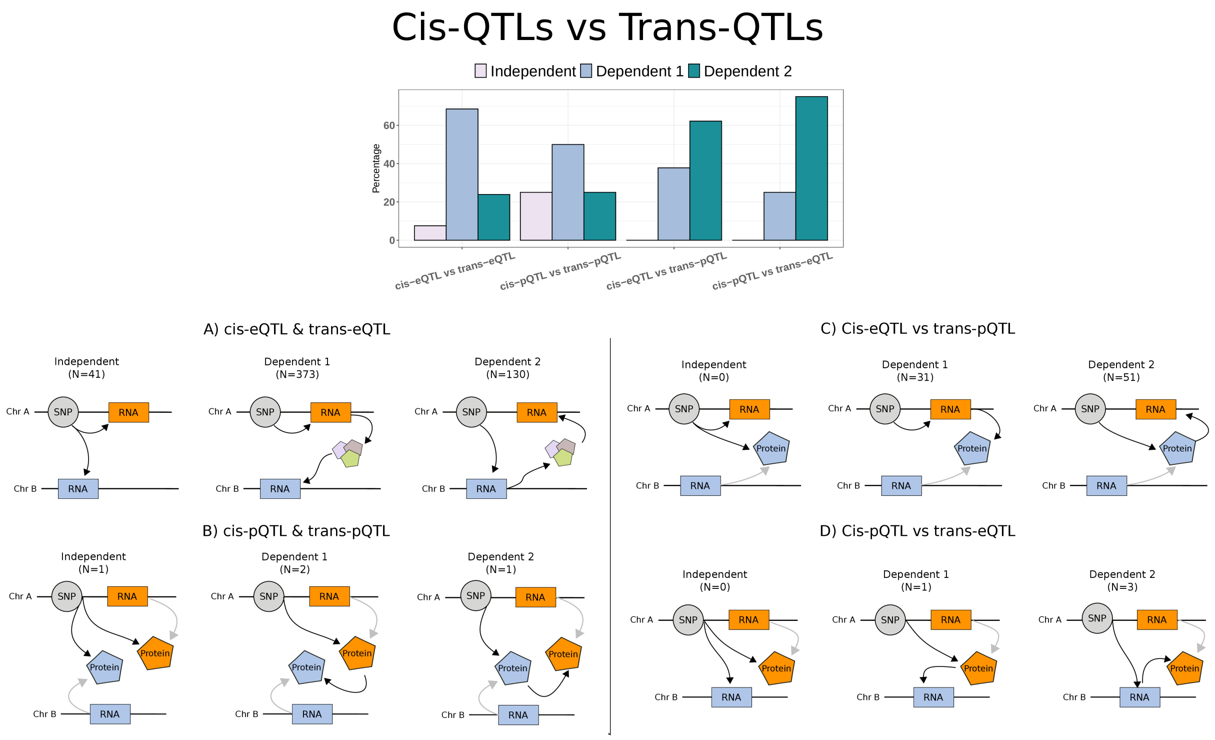


**Supplementary Figure 21 | Causal models for shared cis-trans-QTLs.** The diagram show models used to infer causality for SNPs acting as cis and trans within phenotypes and across different phenotypes. The top barplot summarize the number of models identified and show on the diagrams. Colour code for the different models can be interpreted with the cartoons bellow. **A)** Number of trios of SNP-phenotypes identified for each model when evaluating SNPs with shared cis and trans-eQTLs effects. Dependent model 1 identifies an effect of the SNP on trans expression mediated by a local effect. This is assumed to be mediated by molecules or proteins not measured. Dependent model 2 identify local effects of the SNP in expression were mediated by distal regulation. **B)** Models for SNPs affecting both cis and trans-pQTLs effects. Dependent model 1 identifies an effect of the SNP on distal protein mediated by a the effect on a nearby protein. Dependent model 2 identify local effects of the SNP in protein levels were mediated by distal regulation of other protein. **C)** Models evaluating SNPs with shared cis-eQTL and trans-pQTLs effects. Dependent model 1 identifies an effect of the SNP on a distal protein in mediated by a local effect on gene expression. Dependent model 2 identify local effects of the SNP in expression were mediated by distal regulation of protein levels. **D)** Models evaluating SNPs with shared cis-pQTL and trans-eQTLs effects. Dependent model 1 identifies an effect of the SNP on distal gene expression mediated by a local effect on a protein. Dependent model 2 identify local effects of the SNP in protein abundance were mediated by distal regulation of the expression of other gene.

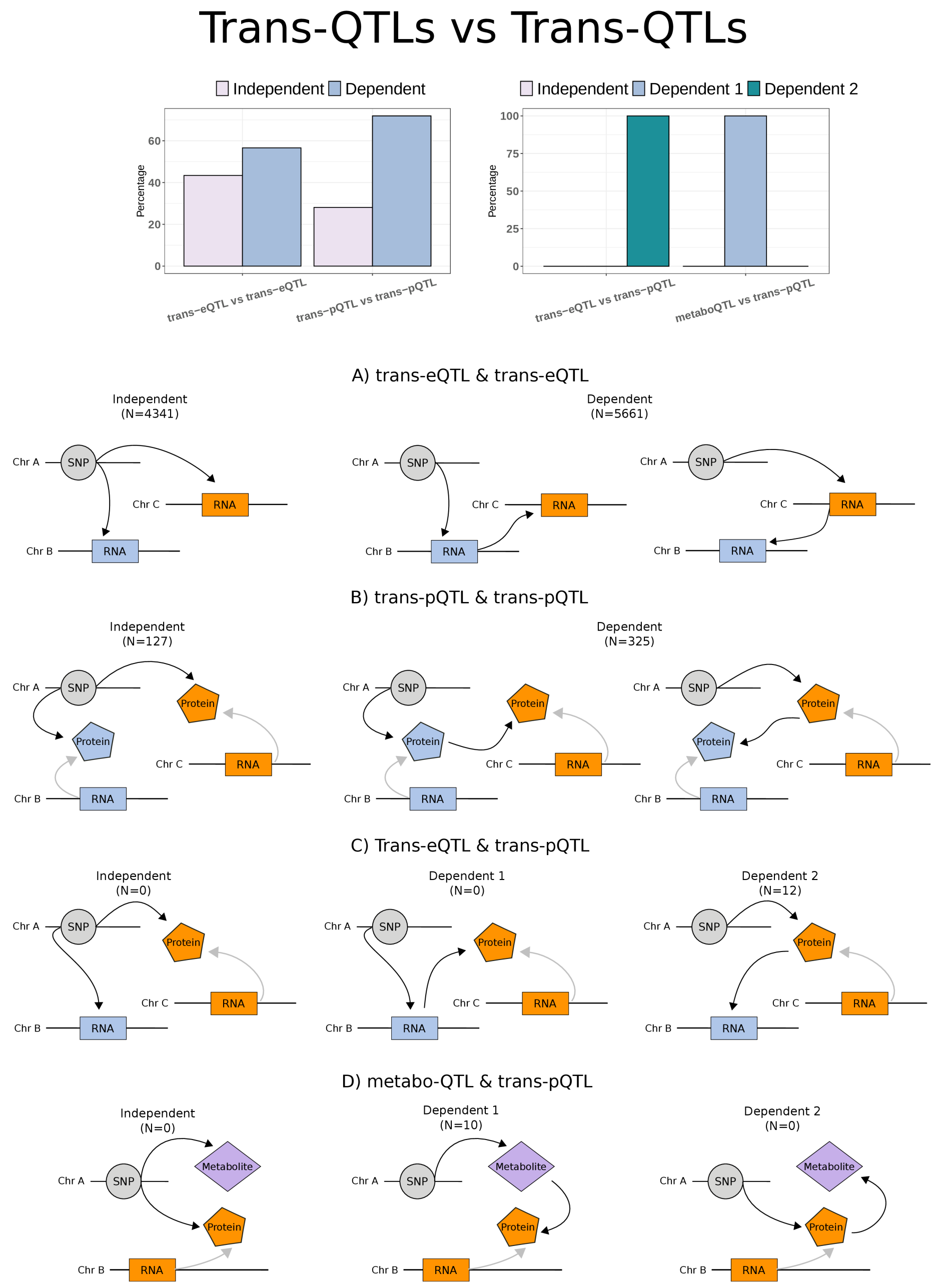


**Supplementary Figure 22 | Causal models for shared SNPs across trans-QTLs.** The diagram show models used to infer causality for SNPs regulating distal phenotypes within and across different phenotypes. The top barplot summarize the number of models identified and show on the diagrams. Colour code for the different models can be interpreted with the cartoons bellow. **A)** Models for shared trans-eQTL effects. **B)** Models for shared trans-pQTL effects. **C)** Models for shared trans-eQTL and trans-pQTLs effects. The only models confidently identify show the effect of a SNP in distal gene expression were mediated by regulation of distal proteins. **D)** Models evaluating SNPs with shared cis-pQTL and metabo-QTLs effects. The only models identify show the effects of the SNPs on distal proteins were mediated by metabolites.

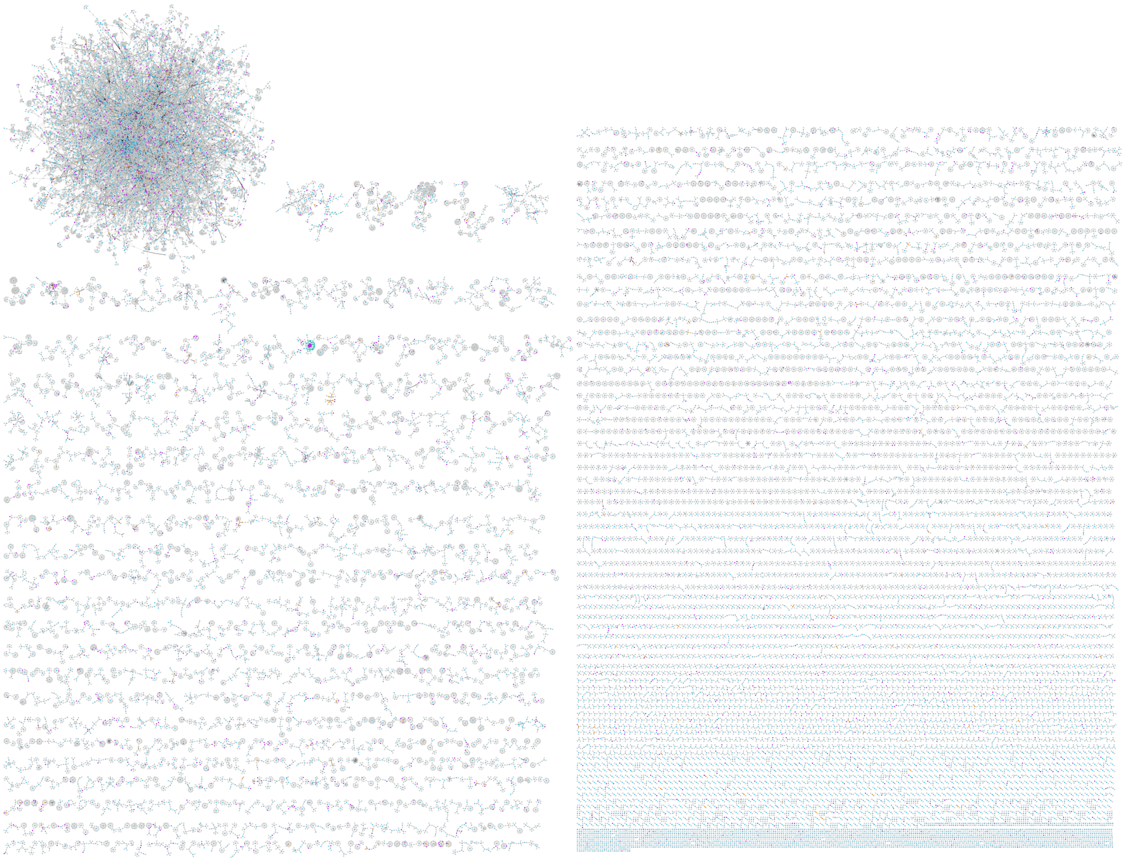

**Supplementary Figure 23 | Full network with all QTLs**. The figure shows the complete network including all QTLs, including 79,733 nodes (15,254 genes, 373 proteins, 172 metabolites and 63,795 SNPs) and 80,645 edges (QTLs), connected in clusters containing between 3 and 19,711 nodes.

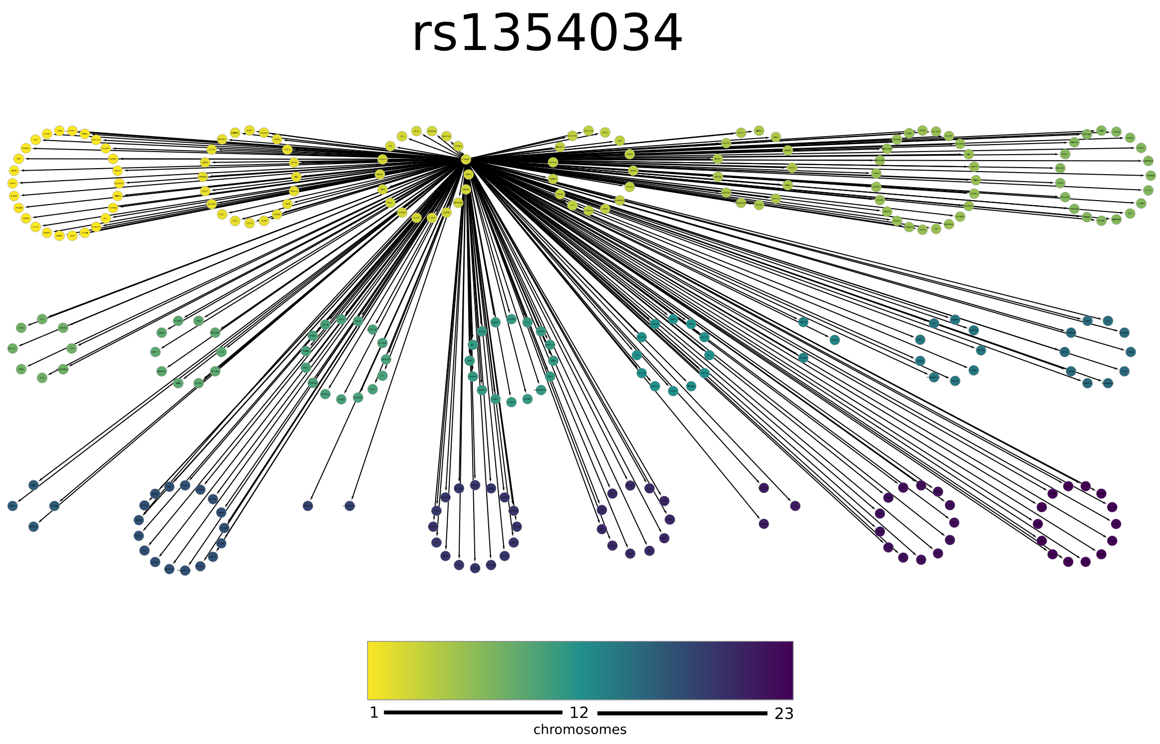


**Supplementary Figure 24 | Trans-eQTLs for rs1354034.** This variant is a trans-eSNP for 297 genes across 23 chromosomes. The diagram shows the 297 genes and the SNP as nodes coloured according to their chromosome location, which are also ordered, with chromosome 1 on the top left, and chromosome 23 on the bottom right. The SNP is located in chromosome 3, and is the origin of all edges.

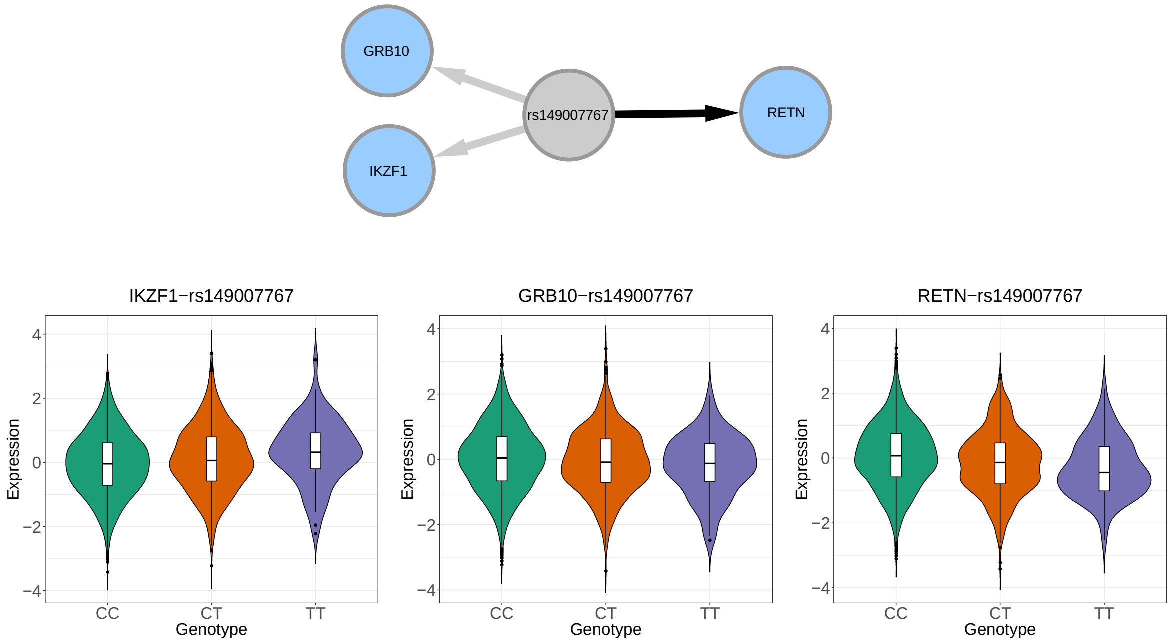


**Supplementary Figure 25 | Cis and trans effects associated to rs149007767**. This variant is a trans-eQTL for *RETN* and show the same direction of effect for two transcription factors in cis: *GRB10* and *IKZF1*, suggesting the trans-eQTL effect may be mediated by one or two of these genes. Causal inference analysis only found supporting evidence for a model where rs149007767 regulation of RETN is mediated by IKZF1 (BIC > 26, compare with the other two possible models for RETN-IKZF1-rs149007767).
